# Reducing Cross-Cultural Comparability Bias Obtained with Measurement Invariance Analysis by Means of Anchoring Vignettes in Heterogeneous Refugee Samples

**DOI:** 10.1101/2023.02.17.23286077

**Authors:** Natalja Menold, Louise Biddle, Hagen von Hermanni, Jasmin Kadel, Kayvan Bozorgmehr

**Affiliations:** Technische Universität Dresden, Dept. of Methods in Empirical Social Research; University Hospital Heidelberg, Section for Health Equity Studies & Migration; Bielefeld University, Dept. of Population Medicine and Health Services Research

**Keywords:** Comparative statistics, measurement invariance, differential item functioning, anchoring vignettes, health system responsiveness, sample heterogeneity, refugees

## Abstract

**Background:** Configural, metric, and scalar measurement invariance have been indicators of bias-free statistical cross-group comparisons, although they are difficult to verify in the data. Low comparability of translated questionnaires or the different use of response formats by respondents might lead to rejection of measurement invariance and point to comparability bias in studies that use different languages. Anchoring vignettes have been proposed as a method to control for the different use of response formats by respondents (RC-DIF) as implemented by means of rating scales. We evaluate the question whether the comparability bias obtained by means of measurement invariance analysis can be reduced by means of anchoring vignettes or by considering socio- demographic heterogeneity as an alternative approach.

**Methods:** We use the Health System Responsiveness (HSR) questionnaire in English and Arabic in a refugee population. We collected survey data in English (n = 183) and Arabic (n=121) in a random sample of refugees in the third largest German federal state. We conducted multiple-sample Confirmatory Factor Analyses (MGCFA) to analyse measurement invariance and compared the results when 1) using rescaled data on the basis of anchoring vignettes (non-parametric approach), 2) including information on DIF from the analyses with anchoring vignettes as covariates (parametric approach) and 3) including socio-demographic covariates.

**Results:** For the HSR, every level of measurement invariance between Arabic and English questionnaires was rejected. Implementing rescaling on the basis of anchoring vignettes provided superior results over the initial MGCFA analysis, since configural, metric and scalar invariance could not be rejected. When using solely socio-demographic covariates, scalar measurement invariance could not be rejected, but configural and metric invariance had to be rejected.

**Conclusions:** Surveys may consider anchoring vignettes as a method to obtain more satisfactory results of measurement invariance analyses; however, socio-demographic information cannot be included in the models as a standalone method. More research on the efficient implementation of anchoring vignettes and further development of methods to incorporate them when modelling measurement invariance is needed.

## 1. Introduction

Cross-cultural social science, as well as comparative psychological, educational, economic and health research has had a longstanding interest in comparisons of persons’ characteristics across or within countries and different subgroups. Self-reports in surveys have been a relevant data collection method. Since the early days of comparative research, ensuring the comparability of data collected on the concepts of interest, for example by means of appropriate translations or data collection methods, has been recognized as a fundamental methodological problem and issue (Harkness et al., 2010). With increasing globalization, but also due to different political systems, religious conflicts and war and poverty, migration and refugee flows are now and will continue in the future to be one of the main human challenges. Conducting surveys in the languages of refugees, i.e. Arabic, and assessing comparability between different refugee languages is therefore a relevant, but under-researched area (Stathopoulou et al., 2019). Our research therefore focuses on the statistical comparability of the measurement of a health related concept between English and Arabic languages in a refugee population.

Information on concepts of interest, such as physical and mental health, well-being, personality, opinions or behaviours have often been collected in surveys by means of multiple indicators (items, questions, manifest variables) that are presented in questionnaires as statements that respondents evaluate with the help of rating scales. Rating scales are graduated response options ordered along a continuum, e.g. ranging from “very bad” to “very good” (example of self-reports and rating scales are provided in Figure 1 and Table 1). Multiple indicators with rating scales or other response options are used with the promise of measuring unobservable concepts of interest, referred to as latent variables, whereas Latent Variable Measurement (LVM) has been a popular statistical measurement approach (Meredith, 1993).

**Figure 1:**
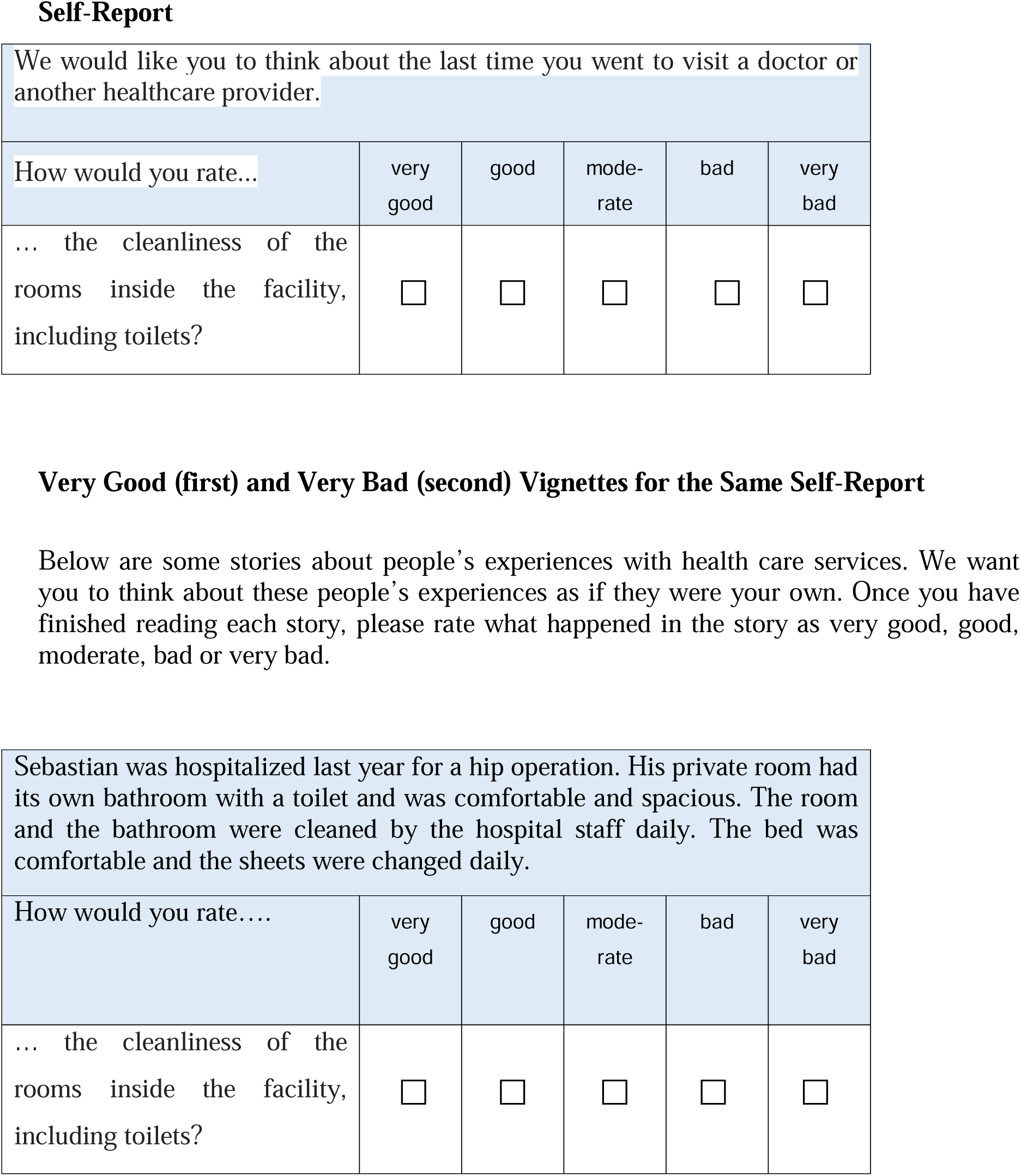
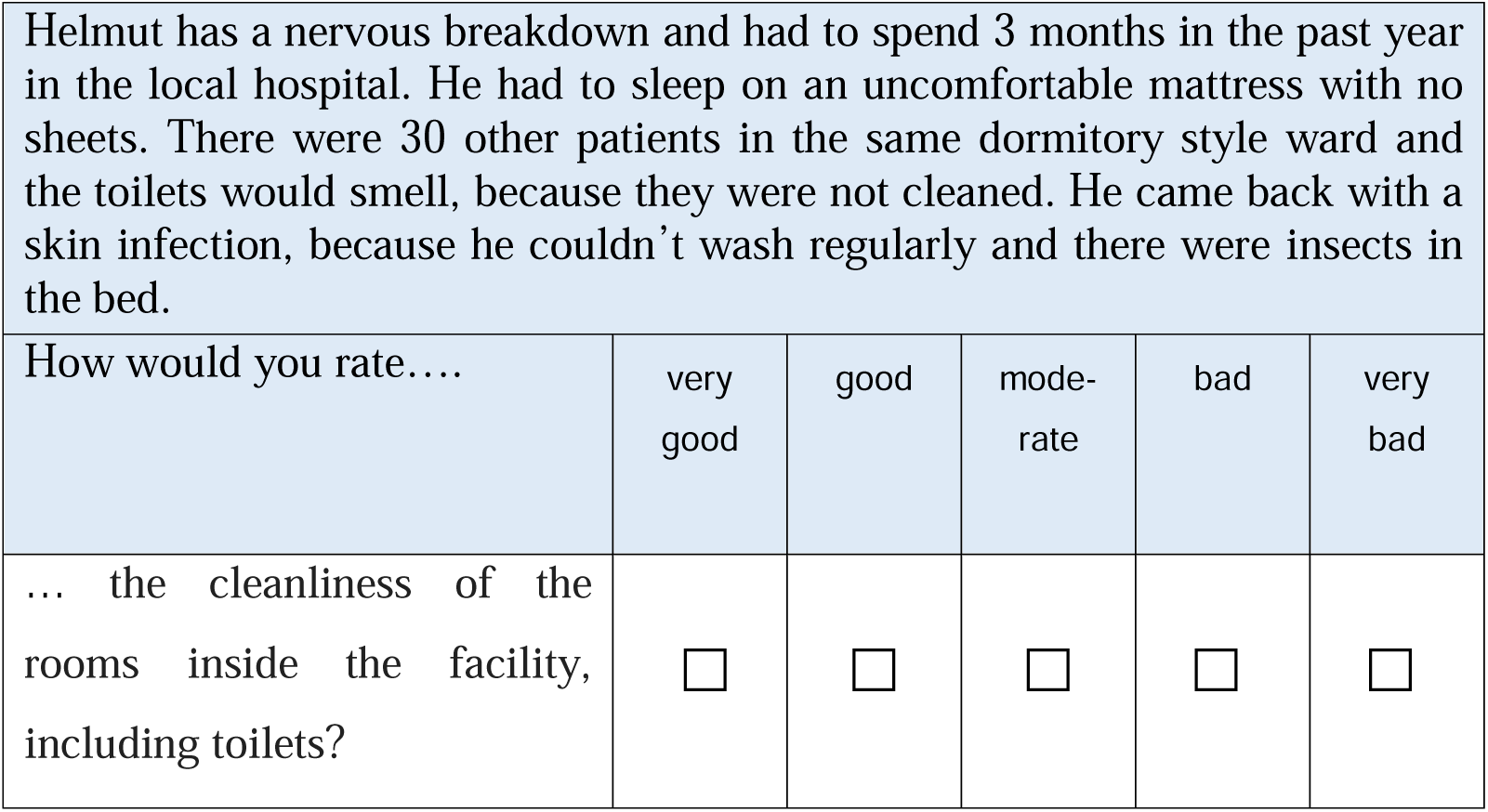
Example for a HSR Self-Report and Anchoring Vignettes for the Two Poles of the Rating Scale (Very Good, Very Bad).

**Table 1.** Indicators of HSR in English questionnaire

| Indicator and its label | Question wording |
| --- | --- |
| attention (time) | ... the amount of time you waited at the doctor's before being attended to? |
| respect (resp) | ... your experience of being greeted and talked to respectfully? |
| communication (comm) | ...the experience of how clearly health care providers explained things to you? (Language and content easy to understand) |
| autonomy (aut) | ... your experience of being involved in making decisions about your treatment? |
| confidentiality (conf) | ... the way health services ensured you could talk privately to health care providers? |
| Choice (choice) | ... the freedom you had to choose your health care provider? |
| quality of amenities (clean) | ... the cleanliness of the rooms inside the facility, including toilets? |

The development of statistical methods such as Confirmatory Factor Analysis (CFA) within the frame of LVM has enabled the presumption of data comparability to be defined and statistically evaluated, typically by means of multiple-sample CFA (MGCFA). Statistical evaluation of potential comparability bias has been referred to as measurement invariance analysis. Measurement invariance means that measurement results are not biased by group membership (Meredith, 1993), i.e. that individuals with identical individual values on the measured concept or variable provide equivalent manifest responses. Measurement Invariance is rejected, not supported or violated, “if individuals from different groups respond to a test item in a dissimilar manner when they are in the same level on the construct being measured” (Kim et al., 2017, p. 524). Measurement invariance analysis has become increasingly popular in empirical cross-cultural research (van de Schoot et al., 2015; Meitinger et al., 2020; Leitgöb et al., 2022), but the results often point to data that is not suitable for the comparisons under investigations, which in turn is associated with data not supporting measurement invariance between different countries and languages (e.g., Davidov et al. 2018; Lee et al., 2020; Zercher et al., 2015; Wu et al., 2007). Whereas numerous studies report the lack of support for measurement invariance in cross-cultural and other group comparisons, little is understood about problems in questionnaires or data collection

situations associated with the comparability bias. It is assumed that the results of measurement invariance analysis can be positively affected if comparability bias is prevented by means of appropriately designed data collection situations or instrumentation (e.g., Leitgöb, 2022; Roberts et al., 2020). Previous research has reported on the lack of support for measurement invariance between different data collection modes, i.e. self- and interviewer administration, in the case of grammar or lexical differences in question wording, or for different realisation of rating scales (cf. Leitgöb et al., 2022 for an overview). This research provides evidence that the data collection situation or data collection instruments are crucial to data comparability. One relevant source of comparability bias may be the ways in which respondents understand and use ratings scales differently. “Seldom” might therefore refer to very different quantities or the same situation might be evaluated as “very good” by one respondent, but as “good” or even “just satisfactory” by other respondents, depending on respondents’ sense of entitlement, experiences, habits, motivations, or activated comparability context.

In the present paper we focus on the potential effect of the different use of rating scales by respondents on the results of measurement invariance analysis and thus on comparability bias in refugee samples. A relevant question has been whether methods that have been used to model the different use of rating scales in other contexts can help to improve the results of measurement invariance analyses, which generally means obtaining better goodness-of-fit statistics when conducting MGCFA analyses. Our first idea is to rely on anchoring vignettes (King et al., 2004; Rice et al., 2012), developed within the frame of Item Response Theory (IRT). Anchoring vignettes have been considered as a method of controlling for the varying use or understanding of rating scales by respondents, referred to in the IRT context as Differential Item Functioning (RC-DIF). RC-DIF means that individuals with the same value on the latent variable have a different probability of choosing the corresponding answer (Holland and Wainer, 1993). RC-DIF can be thought of as the opposite of measurement invariance, as in the presence of RC-DIF measurement invariance might not be supported in the data. Anchoring vignettes are situation descriptions that correspond to a response category (Figure 1). “Vignettes represent hypothetical descriptions of fixed levels of a construct… and individuals are asked to evaluate these in the same way that they are asked to evaluate their own experiences…” (Valentine et al., 2009, p. 175). In questionnaires, respondents provide both their self-evaluations and evaluations of anchoring vignettes. For the Health System Responsiveness (HRS, Valentine, 2009, see Figure 1, Table 1 and 2) –the concept we use in the present study – respondents evaluate their own most recent experiences with health care institutions,e.g. timeliness of the last visit to a doctor, as “the amount of time you waited at the doctor’s before being attended to” on a rating scale consisting of “very good”, “good”, “moderate”, “bad”, “very bad”. In addition to this self-report, respondents evaluate vignettes which are descriptions of fictive situations a person experiences during a visit to the doctor, i.e. waiting for hours in the case of a “very bad” vignette. The RC-DIF is given, if respondents tend to evaluate the vignettes inconsistently with the described level of the concept. This is the case if, for example, the “very bad” vignette situation is evaluated with “bad”, “moderately” “good” or “very good” by respondents. The control of or adjustment for the RC-DIF means that information on responses to vignettes is used in the regression models with self-reports. Anchoring vignettes are therefore promising in reducing comparability bias and are becoming increasingly popular. They have been implemented in some large-scale international surveys, i.e. Survey of Health, Ageing and Retirement in Europe (SHARE, d’Uva & Donnell, 2011), Programme for International Student Assessment (PISA, Marksteiner et al., 2019), Wisconsin longitudinal Study (WLS; Grol-Prokopczyk, Freese, & Hauser, 2011), or World Health Survey (Valentine, 2009). The HSR instrument for self-reports and anchoring vignettes we use in our study (Valentine, 2009, see Figure 1 and Tables 1, 2) are taken from the World Health Survey (WHS).

**Table 2.**
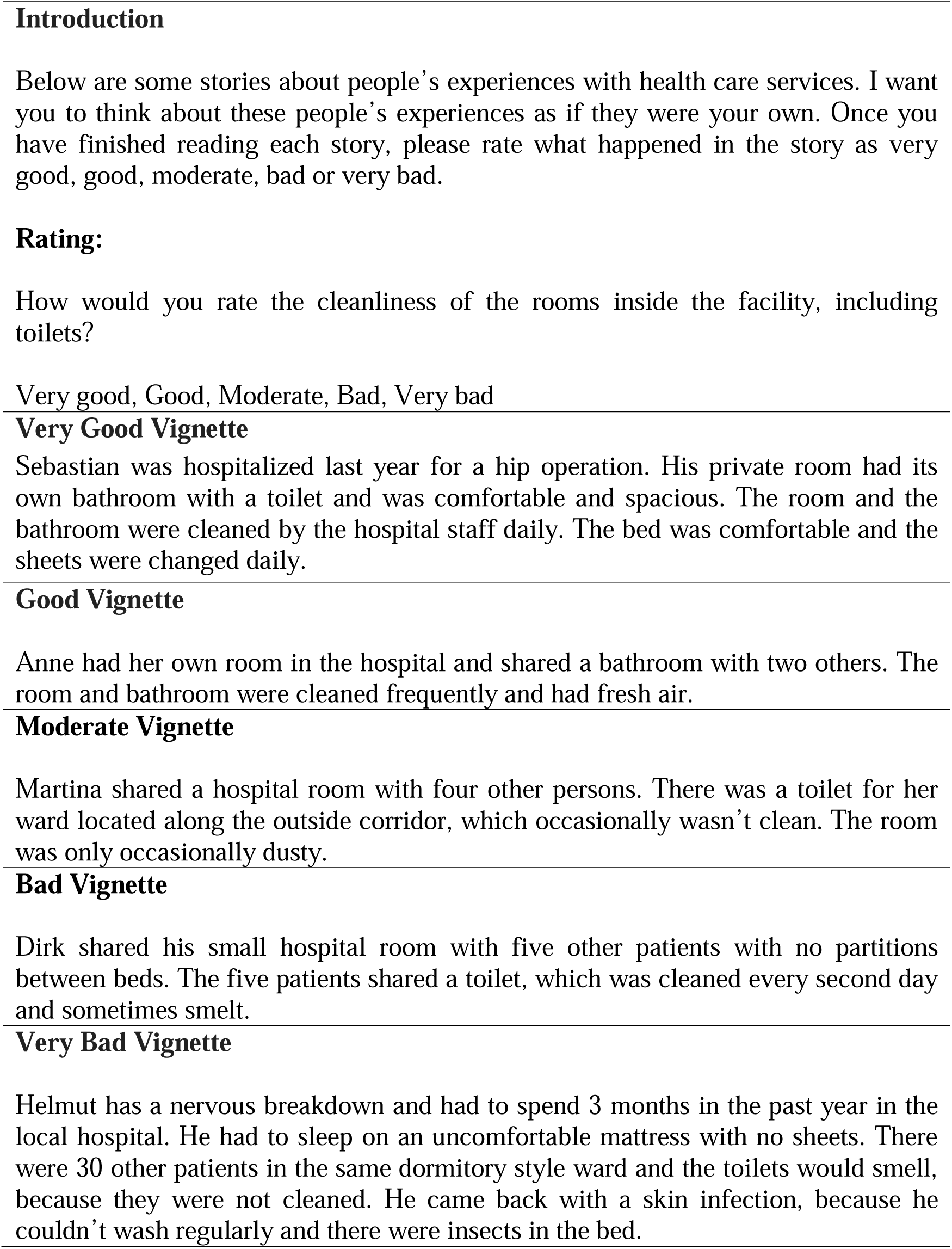
Survey Question and Anchoring Vignettes of the HSR indicator “Quality of Basic Amenities”

Of course, if anchoring vignettes are a means of controlling or adjusting the data for RC-DIF, their use can also influence the results of measurement invariance analysis by means of MGCFA. He et al. (2017) and Marksteiner et al. (2019) demonstrated a more satisfactory model fit of measurement invariance analyses when data was rescaled using vignette evaluations in PISA (rescaling procedure is introduced in 1.2). However, the results of these studies are mixed and more research is needed, especially when it comes to health-related topics and refugee populations. The implementation of anchoring vignettes also requires that additional information is asked for in questionnaires. This would be burdensome, particularly if a full set of vignettes (e.g., five in the case of five category rating scales) is used for each indicator. For the seven indicators of the HSR short scale, 35 vignettes should be additionally included in the questionnaire. This limits the use of vignettes in the survey practice and makes alternatives relevant. Hox et al. (2015) use demographic information as covariate variables in the MGCFA measurement invariance analysis when comparing different modes of data collection. In a similar way, we evaluate whether the use of demographic information in MGCFA models can help to reduce comparability bias.

Our research addresses the use of anchoring vignettes and socio-demographic information to obtain more satisfactory results of measurement invariance analysis. We use data collected in Germany on HSR in English and Arabic speaking refugee samples. Extending on previous research (He et al., 2017; Marksteiner et al., 2019), we address a health research topic among refugee populations. In doing so, we replicate the studies by He et al. (2017) and Marksteiner et al. (2019). We additionally consider MGCFA covariate models incorporating information on RC-DIF predicted from vignettes’ ratings. Further, we consider demographic variables without information on RC-DIF from vignettes’ ratings.

Our paper is structured as follows. We firstly provide specifications of measurement invariance models and a research overview of their use in cross-cultural research a (section 1.1). Next, we provide specifications for a parametric and non-parametric approach to the use of anchoring vignettes to model RC-DIF (section 1.2). We specify our research questions on this basis (section 1.3). In section 2 we describe the study, data and materials as well as data analysis method. In section 3 we provide the results. Discussion and conclusion are provided in sections 4 and 5.

### 1.1 Measurement Invariance

Measurement invariance analysis provides information on whether between-group comparisons of latent variables or summarized scores deliver valid results, as certain levels of measurement invariance are necessary in order to make bias free statistical comparisons (Millsap, 2011; Meredith 1993; Wu et al., 2007). Measurement invariance analysis is typically conducted by a sequence of steps of MGCFA.

In MGCFA, a measurement model (that is a CFA model) is evaluated for observed scores *Y* on an indicator of individual *i* within group *j*:

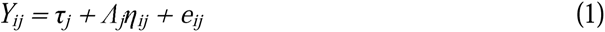

where τ_*j*_ represent intercepts and Λ*j* represent factor loadings for the group *j*, and η*_ij_* and *e_ij_* represent common scores and residuals for the individual *i* in group *j*, respectively.

The following increasing degrees (or levels) of measurement invariance are relevant to the survey context (Hox et al., 2015; Gregorich, 2006)^1^, with each subsequent one including the preceding (Meredith, 1993; Millsap, 2011):

(a) Configural invariance is defined in equation (1) and holds when the number of factors (latent variables) and indicators per factor are comparable across groups. If the configural invariance holds (that is, not rejected by the data), however, statistical comparisons of latent variables or simple sum scores are not sensible among groups.
(b) Metric or weak invariance holds, if Λ*_j_* = Λ for all groups, that is, if loadings that reflect the strength of associations between the manifest and latent variables are comparable among groups. Metric invariance should be given to compare correlations among groups. To evaluate metric invariance, equality constraints on factor loadings among the groups are introduced into the configural model. Equality of factor loadings and therefore the presence of metric invariance is proven if the introduction of the restriction does not significantly decrease model fit.
(c) Finally, scalar or strong invariance among groups holds, if τ*_j_* = τ invariance is evaluated by restricting the intercepts of the manifest variables to make them equal among groups. Again, this restriction should not significantly decrease model fit. Satisfying scalar invariance allows for valid comparisons of both latent mean scores and means of summarized scores.

This description of different degrees of measurement invariance shows, therefore, that weak and strong measurement invariance are prerequisites of the bias-free cross-group comparisons, as their violation means that results are confounded with the group comparability bias in the measurement.

With respect to measurement invariance in cross-cultural studies, researchers often fail to support strong or even weak invariance in their data, as shown by Wu et al. (2007) for the Trends in International Mathematics and Science Study (TIMMS), by Davidov and colleagues (Davidov et al., 2008, 2014, 2018; Zercher et al., 2015) for different concepts of the European Social Survey (ESS), or by Meitinger (2017) for some of the concepts in the International Social Survey Program (ISSP). Dong and Dumas (2020) report in a meta-analysis that scalar invariance between ethnic groups was not supported for any of the personality inventories considered. One line of research tried to develop less restrictive data analysis methods (Muthén & Asparouhov, 2013; Asparouchov & Muthén, 2014), while the other line of research has been targeting the question as to which circumstances of data collection situation or cognitive respondents’ problems are associated with the rejection of statistical measurement invariance (Leitgöb, 2022; Roberts et al., 2020).

Differences in response behaviour can be systematically described using the theory of the cognitive response process by Tourangeau et al. (2000) that comprises four separate steps when answering a survey item: The comprehension of a survey question, information retrieval, judgement, and finally, response according to the given response options. When using rating scales, cross-cultural differences in response behaviour during the last step of the cognitive response process would manifest in response styles or response sets, such as acquiescence (Paulhus, 1991), or middle and extreme response tendencies (e.g., van Vaerenbergh & Thomas, 2013). Previous research identified cross-cultural differences in response tendencies depending on education, acculturation, or Hofstede’s dimensions of individualism, power distance or masculinity (Yang et al., 2010). Response styles and response sets may bias the data and limit their comparability, with the manifestations in rejecting measurement invariance in the corresponding statistical models.

Knowing such sources sheds more light on the sensitivity of the measurement invariance modeling and practical significance of its results. Rejection of metric invariance, for instance, would imply that extreme response style is present in the data (Kline, 2016; Gregorich, 2006; Cheung & Rensvold, 2002). Rejecting scalar invariance would point to the presence of additive systematic measurement error, such as acquiescence (Kline, 2016; Cheung & Rensvold, 2002). Research on rating scales has shown that use of different numbers of categories or different category labelling in independent respondents’ groups lead to the rejection of metric and scalar invariance (Menold & Kemper, 2015; Menold & Tausch, 2016).

### 1.2 Modelling and Controlling RC-DIF by means of Anchoring Vignettes

Data can be adjusted using anchoring vignettes when a parametric or a non-parametric approach is implemented (King et al., 2004; King & Wand, 2007). In case of a non-parametric approach, vignette assessments (*z*) are used to rescale the self-assessments (*y*). *J* is the notation for the number of vignettes (*j* = 1, …., J). The rescaling produces a new variable *C* as follows (Equation 2; King et al., 2004; King & Wand, 2007):

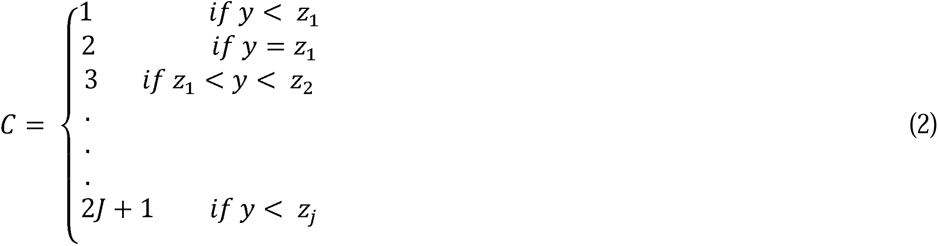

The use of rescaled data for measurement invariance analysis is referred to in the following as non-parametric data adjustment.

The parametric approach, on the other hand, uses a hierarchical ordered regression model (abbreviated CHOPIT) to predict respondents’ self-assessment (*s*) by their evaluation of vignettes (*v*) (King et al., 2004; King & Wand, 2007; Tandon et al., 2003). In this approach, a respondent (denoted by *i* = 1, …, N) has an unobserved level 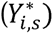 of his/her self-assessments (*s* = 1, …., S), given the actual observed level of self- evaluation (*µ_i_*), as shown in Equation 3.

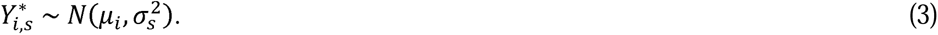

The actual level *µ_i_* is a linear function of observed covariates *Xi* (e.g. gender, age, education), see equation 4.

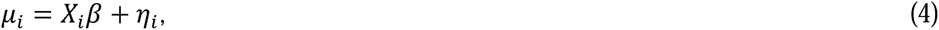

where *β* is the parameter associated with the impact of covariates and *η* the normal random effect.

The reported survey response *y_i,s_* is also dependent on the chosen response category *k* (k = 1,…, Ks) as follows:

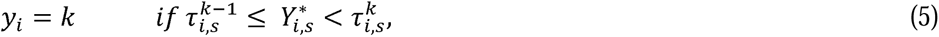

where *τ_i_* is a vector of ordered thresholds (ranging from -∞ to +∞). The thresholds are defined as follows (equation 5):

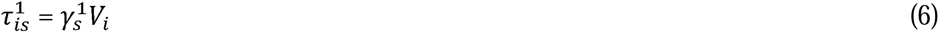

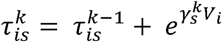 where *Vi* is a vector of covariates and 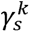 a vector of unknown threshold parameters.

For the vignettes, there is also a predicted value for each respondent from the observed vignette value θ*j*, while respondents are denoted with *l*:

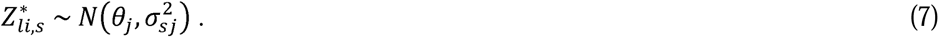

The observed vignette values (*z*) depend on response categories as follows:

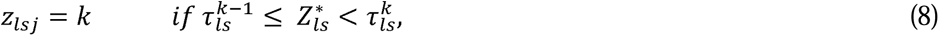

Correspondingly, the values of vignette thresholds are predicted as follows:

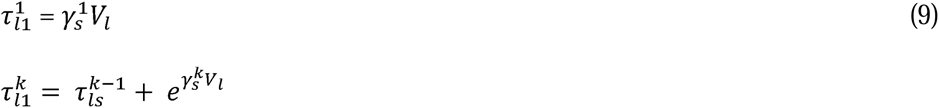

In both self-reports and vignette components, the thresholds vary on the same covariate variable components (*Xi* vs *Vi*). The CHOPIT model estimates in parallel the self-component (mean location of self-assessments), the vignette component (mean location of the vignettes) and thresholds on the basis of vignettes. The use of estimates for vignette components in other models is referred to in the following as parametric adjustment by means of anchoring vignettes.

One line of research on anchoring vignettes addresses the possibility of evaluating the general assumptions of their use, namely vignette consistency and vignette equivalence (e.g., d’Uva et al., 2011; van Soest et al., 2011). Vignette consistency assumes that response behaviour is the same in the case of vignette evaluations and self-assessments, while vignette equivalence means that the same latent dimension explains the responses to all vignettes. The equivalence needs to hold not just within the vignette set, but – in light of response consistency – also between vignettes and self-report questions (Hopkins and King, 2010; Salomon et al., 2004; Greene et al., 2021). Research has particularly evaluated vignette consistency when using correlations with third variables, and the use of objective measures for these variables has been suggested as the best solution (King et al., 2004; van Soest et al., 2011, d’Uva et al., 2011). The results from fulfilment of these general assumptions have been mixed, however (see Greene et al., 2021 for an overview).

Research has also been conducted on the usability of vignettes to actually improve the comparability of data (i.e. adjust for the RC-DIF). One relevant finding is that adjustments with vignettes were associated with a higher criterion validity. King et al. (2004) showed this for visual ability, van Soest et al. (2011) for drinking behaviour and Mõttus et al. (2012) for a personality measure. However, He et al. (2017) obtained mixed results with respect to validity coefficients. Marksteiner et al. (2019) found a higher internal consistency of rescaled data when using the non-parametric rescaling for non-cognitive skills of students in PISA.

The vignettes’ effect on RC-DIF and adjustment of data for comparability – the specific aim of the vignette approach – has been mainly evaluated by comparing adjusted and non-adjusted results (both, parametric modelling and non-parametric rescaling), obtaining more plausible conclusions when using anchoring vignettes (e.g., Kyllonen & Bertling, 2014; Rice et al., 2012; Mõttus et al., 2012). However, such a comparison does not allow for a statistical test and therefore does not provide strong evidence that anchoring vignettes reduces the comparability bias. By way of contrast, measurement invariance analysis (as described in the previous section) allows the suitability of data for statistical comparisons to be tested directly. The research that applies MGCFA models on rescaled data (with non-parametric rescaling, Eq. 2) is available for PISA. He et al. (2017) found a slightly reduced difference in the model fit when evaluating metric invariance. The authors also found the inconsistent use of anchoring vignettes to be correlated with low socio-economic status and low cognitive skills, which point to the relevance of these factors for comparability bias. Marksteiner et al. (2019) also used PISA data on non-cognitive skills and found a higher level of measurement invariance for rescaled data (non-parametric rescaling, Eq. 2) for some contents, but not for others. The authors conclude that the effect of rescaling on the basis of anchoring vignettes on the results of measurement invariance may be dependent on the topic. They also suggest further research when using parametric approach.

### 1.3 Research Questions

So far, we can state that on the one hand, research has often found a comparability bias in large-scale surveys such that strong or even weak measurement invariance are rejected in the data. On the other hand, anchoring vignettes have been used as an approach of control of RC-DIF and it can be expected that information on RC-DIF from anchoring vignettes is utilisable for measurement invariance analysis. Previous research (He et al., 2017; Marksteiner et al., 2019) supported this assumption for PISA data in educational research. We extend previous research by addressing both, a health topic and a specific refugee population, further implementing the parametric modelling. As outlined earlier, the parametric approach makes particular use of socio-demographic and other respondents’ background variables (covariates, see equations 6, 9). The administration of anchoring vignettes may depend on cognitive skills (He et al., 2017) and response styles, and the latter were found to be dependent on socio-demographic variables (Yang et al., 2010). In other contexts, consideration of socio-demographic variables helped in supporting assumptions of measurement invariance, i.e. when evaluating mode effects in non-experimental data (Hox et al., 2015). Therefore, when applying the parametric approach, the potential effect on measurement invariance can be due to both, sociodemographic information and anchoring vignettes, which should be separated from each other. This also has practical consequences, if comparable results with respect to measurement invariance are obtained incorporating socio-demographic information. If so, socio-demographic information can be used to control for RC-DIF thereby avoiding the workload associated with anchoring vignettes.

With this in mind, we address the following research questions: How does information on RC-DIF obtained from anchoring vignettes alter the results of measurement invariance analysis? Does implementing non-parametric rescaling and incorporating CHOPIT- predictions into the analysis of measurement invariance provide similar results with respect to configural, metric and scalar invariance? Are these results comparable to those that consider solely socio-demographic covariates only?

## 2. Methods

### 2.1 Data

This analysis uses data from a population-based, cross-sectional survey among refugees living in collective accommodation centres in the German state of Baden-Württemberg, conducted as part of the RESPOND project *(‘Improving regional health system responses to the challenges of migration through tailored interventions for asylum-seekers and refugees’* – RESPOND) from February to June 2018. The development of the questionnaire, and the sampling and data collection approach have been described in detail elsewhere (Biddle et al., 2019; Biddle et al., 2021). The pen and paper questionnaire comprised established instruments covering health status, utilization of health services, HSR (incl. corresponding anchoring vignettes), as well as several socio- demographic characteristics. It was developed in German and English and translated into the refugee languages (among others into Arabic, which is relevant to this paper) using a team approach (Behr, 2009). The questionnaire was subsequently assessed in the form of a cognitive pretest and refined accordingly (Hadler et al., 2017).

Sampling of participants was conducted on the basis of residential units which included initial reception and regional accommodation centres as no population-based registry of all asylum seekers in the state was available for research purposes. A two-stage sampling design was employed for initial reception centres: First, six of nine centres were purposely selected based on their size, geographical location and administrative responsibility. Second, 25% of rooms (depending on their occupation status) were selected. For regional accommodation facilities, a record of all 1938 facilities in the state was compiled and a random sample of 65 facilities drawn, balancing the number of refugees in each accommodation facility. All individuals living in the selected rooms (reception facilities) or facilities (regional accommodation centres) who could speak one of the study languages and were 18 years or older were invited to participate.

Data was collected by trained, multilingual staff visiting each selected accommodation facility on two consecutive days. Eligible individuals were approached in person by the research staff, who explained the purpose of the study with the aid of pre-recorded audio-messages where there were language barriers. The staff distributed information leaflets, the questionnaires as well as non-monetary, unconditional incentives. Participants could either return the questionnaire to the research staff in person or by post in a stamped envelope. All methods were carried out in accordance with relevant guidelines and regulations, such as ethical standards and the data protection regulations of the European Union (GDPR). All persons were provided with detailed information on the purpose and content of the study, voluntary participation, data collection purpose, data handling and participants’ rights. Informed consent was obtained from all study participants.

Out of 1429 eligible individuals, 1201 were invited to participate in the study. A total of 560 participants completed the survey (reception centres: 149; accommodation centres: 411), with a total response rate of 39.2%. This response rate is satisfactory due to decreased participation rates in surveys, while response rates of 30% or lower are rather usual (Harrisson et al., 2019; Meyer et al., 2015). Since anchoring vignettes for HSR are implemented in English and Arabic only, the analyses are necessarily restricted to these two groups. Of those respondents who used English to participate in the study (n = 183), 27% were from Gambia, 43% from Nigeria, 6% from Sri Lanka and 16% had other countries of origin that were not specified further in the questionnaire. Of the Arabic speaking persons (n = 121), 56% were from Syria, 26% from Iraq and 14% of other origin. Table 3 provides further information on the socio-demographic characteristics of our sample (N = 304).

**Table 3.** Summary of Sample Characteristics and Selected Socio-Demographic Variables

|  | share<br>in % | n |
| --- | --- | --- |
| English speaking | 60 | 183 |
| Arabic speaking | 40 | 121 |
| Female | 23 | 69 |
| Age 26-30 | 19 | 59 |
| Age 31-35 | 15 | 45 |
| Age 36-40 | 9 | 28 |
| Age 41+ | 11 | 33 |
| Insurance electronic Card yes | 38 | 114 |
| Insurance electronic Card – missing data | 6 | 19 |
| Education no school / do not know | 25 | 75 |
| Education mandatory school | 19 | 59 |
| Education high school | 35 | 105 |

Note. $N = 304$ (100%) total English and Arabic speaking persons; $N = 245$ who administered HSR and vignettes

### 2.2 Material

HSR is defined as “aspects of the way individuals are treated and the environment in which they are treated during health system interactions” (Valentine et al., 2009, p. 138). The inventory aims to measure the latent concept of the non-technical quality of care received during healthcare interactions, including respectful and confidential treatment by health care personnel, clarity of communication and information, timeliness of treatment and the quality of basic amenities. HSR was first implemented in the WHO Multi-Country Survey Study and subsequently embedded in the World Health Survey (WHS), collecting data in over 70 countries. It is currently part of the WHO Study on global ageing and adult health (SAGE). However, it has not previously been used specifically in refugee populations (Mirzoev & Kane, 2017) and no analysis of measurement invariance for HSR has previously been available. HSR utilizes a five-category, fully verbalized rating scale ranging from “very bad“ to “very good“ (Table 1). In addition, anchoring vignettes for the HSR are used in WHS, making HSR particularly relevant for the aim of our study (Table 2).

We used the short-form version of HSR included in the WHS, restricting our questions to ambulatory care only (see Table 1 for the question wording). The HSR instrument as implemented in the WHS demonstrated moderate test-retest reliability (kappa values of 0.40-0.49 across domains) and border internal consistence (Cronbach’s alpha of 0.65) (Valentine et al., 2009).

Prior to data collection, the translated version of the HSR instrument was included in a cognitive pretest (Hadler et al., 2017). Using probing and think-aloud techniques, these pretests evaluated the intelligibility of the items and assessed potential unintended misunderstandings with nine refugees in five languages, including English and Arabic. This pretest resulted in the simplification of the question format and clarifications of particular terms used. The reliability of the improved HSR as a latent dimension was sufficiently high in the whole asylum seekers sample (factor analysis based *ro* = .87 (Raykov & Marcoulides, 2011); all loadings were higher than .50).

Respondents evaluated vignettes in addition to self-assessment on HSR (see Table 2 for an example of vignettes). Using vignettes for each of seven indicators and each response category resulted in 35 vignettes, which means that an additional 35 questions had to be included in the questionnaire. To reduce the workload of respondents and due to the limited number of questions that could be included in the survey, we used five different sets of vignettes in each language, with each set being randomly assigned to a respondent group. The first set of 21 vignettes contained the top, the middle and the bottom vignettes for each of the HSR indicators. The other four sets included five sets of vignettes for each response category for two or one of the indicators of HSR (set two attention and respect, set three communication and quality of amenities; set four confidentiality and choice; set five autonomy).

### 2.3 Data Analysis

For the HSR, we conducted an MGCFA analysis. For all MGCFA analyses we used the software Mplus 8.7. To compare loadings and intercepts, the factor means and variances were set to 1 and estimated freely (cf. e.g. Byrne, 2016; Kline, 2016). To account for ordinality and non-normality of data we used Robust Maximum Likelihood estimator (MLR; Muthén and Muthén, 2014). In the case of ordinal data with five to seven categories and small samples, MLR method provides more stable and valid results than use of estimators for ordinal data (Li, 2016). To validate the results, we also conducted analyses for ordinal (ordered categorical data).

The model fit of MGCFAs was evaluated using the chi-square test (CMIN), the Root-Mean-Square Error of Approximation (RMSEA), and the Comparative Fit Index (CFI) (Beauducel & Wittmann, 2005). The CFI should be 0.95 or higher, while the RMSEA of 0.08 or less indicates an acceptable fit (Hu & Bentler, 1999). A significant change of CMIN (Meredith, 1993) or a change of ΔCFI ≥.005 and ΔRMSEA ≥.010 indicate significant differences in model fit if the samples are small (n < 300) and unequal (Chen, 2007), thus demonstrating a lack of measurement invariance. To compare the different unnested models with different covariate variables and those with different sample sizes, sample size adjusted Bayesian Information Criterion (BIC) was used, where lower values indicate a better model fit and a change of BIC ≥ 6 indicates significant change (Raftery, 1995).

#### Measurement Invariance Analysis with Data Rescaling (Non-Parametric Approach)

For the non-parametric approach, we used the rescaling procedure for each of the HSR indicators as introduced in equation 2. Because the non-parametric approach requires each respondent to evaluate vignettes (while a selection of vignettes can be used, i.e. top or bottom or top, middle, bottom, see King et al., 2004 and He et al., 2017 respectively), we calculated means for top, middle and bottom vignettes from all respondents’ groups (see information on different groups using different vignette sets above). The analysis to rescale the self-reports on HSR indicators was conducted using R (see equation 1 as well as Wand and King (2007) for details; the software source is included in **Appendix 1**). A similar procedure, also using two or three vignettes (top, middle, bottom) was implemented in He et al. (2017) and Marksteiner et al. (2019). The anchor package of R accounts for the misordering of vignettes and predicts the lowest (Cs) and highest possible (Ce) ratings for each of the HSR items. Similar to He et al. (2017) and Marksteiner et al. (2019), we considered both predictions. The rescaled variables were subsequently used in MGCFA analyses to evaluate measurement invariance.

#### Measurement Invariance Analysis with Covariates

In addition, we evaluated the RC-DIF by means of the parametric approach (Equations 3 to 8, section 1.2), using glamm function of Stata (Grol-Prokopczyk et al., 2011; Rabe-Hesketh and Skrondal, 2002; 2003, **Appendix 2**). Data from the groups who used vignette sets 2 to 5 was included. In the CHOPIT analysis, the covariates were language, gender, age, and having an electronic health insurance card. We considered these variables to be relevant to health care experiences. With respect to other variables, such as economic, occupational status or living conditions, the respondents were deemed to be too similar due to their status as asylum seekers and their current stay in refugee centres. Also, our small sample size prohibited the use of too many predictors. As the education variable had a reasonable number of missing values (n = 21 English and n = 15 Arabic), we excluded it from the CHOPIT analysis to avoid a substantial decrease of sample size. To be able to use the vignette data in the MGCFA, we saved the predicted threshold parameters (Equation 8, see glamm code in **Appendix 2**; Grol-Prokopczyk et al., 2011; Rabe-Hesketh and Skrondal, 2002). The results of the CHOPIT analysis for an example indicator of the HSR can be found in **Supplementary Material**, as these are out of our focus and the procedure was merely used as an auxiliary step to predict thresholds for subsequent use in the measurement invariance analysis.

There are no solutions in the literature for the implementation of corrections when relying on the parametric approach within anchoring vignette research. However, within LVM in Structural Equation Modeling, one method of controlling for sample heterogeneity has been to include covariate variables in the analysis (Lubke & Muthén, 2007; Muthén 2002). Therefore, in the MGCFA model of HSR we included predicted threshold parameters as covariates which were regressed on observed indicators. So, in our models, the variation in observed indicators is explained by both the latent variable and by vector of covariates. Such models have been referred to as covariate models within CFA (Lubke & Muthén, 2007). To define our model we extend the equation (1) as

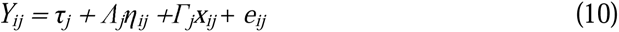

where *x* are covariates and Γ*j* are the regression weights. The covariates are either the predicted thresholds from the CHOPIT analysis or socio-demographic variables. We did not postulate a second latent variable to explain covariates, since the socio-demographic variables are not expected to build a latent variable. Similarly, the response thresholds predicted from the vignettes can be explained by a latent variable if vignette equivalence holds. To overcome this assumption, we considered predicted indicator thresholds as manifest covariates (Equation 10). Overall, CHOPIT predicted four thresholds for each of seven indicators of HSR, which means having a vector of 28 covariates in equation 10, which challenges the model complexity and small sample sizes we had. We therefore included predicted thresholds with a significant path on at least one indicator of the HSR. The resulting covariate model is shown in Figure 4 and the software code is provided in **Appendix 3**. The covariate MGCFA model with socio-demographic variables is shown in Figure 5; see **Appendix 4** for the Mplus code.

## 3. Results

### 3.1 Vignettes Accuracy

The proportion of accurately ordered vignettes was 68% in the English speaking group and 76% in the Arabic speaking group. It can be seen from Figure 1 that vignettes were evaluated similarly by the two language groups, and according to a MANCOVA (Multivariate Analysis of Covariance) there were no significant differences between the mean evaluation of bottom, moderate and top vignettes (*Pillai-Spur(PS)* = .01; *F*(3,228) = 1.04, *p* > .10, η = 0.01). There were no significant differences in the evaluation of vignettes between men and women (*PS* = .02; *F*(3,217) = 1.26, *p* > .10, η = 0.02), or between different age groups (*PS* = .07; *F*(9,636) = 1.57, *p* > .10, η = 0.02). However, respondents with higher education ranked the top (*M* = 4.23; *SD* = 0.82) and bottom (*M* = 1.97, *SD* = 1.1) vignettes more consistently (*PS* = 0.10; *F(6, 424)* = 3.61, *p* < .01, η = 0.02). Finally, respondents without electronic health insurance cards evaluated the “very good” (top) vignette closer to its rank 5 (*M* = 4.16, *SD* = 0.91) than other respondents (univariate effect F(1,230) = 3.84, *p* = 0.05, η = 0.02).

### 3.2 Measurement Invariance Initial Model of HSR

Figure 2 shows the MGCFA model and Tables 4 and 5 provide an overview of estimated parameters for the configural model of HSR, while Table 6 provides goodness of fit statistics. The configural invariance was not supported, because the configural model was associated with a pure model fit due to the significant CMIN, the RMSEA reasonably above and the CFI far below the benchmark (Table 6). To obtain an acceptable baseline model to evaluate the comparability of loadings and intercepts, we inspected misspecifications looking at the Modification Indexes (MIs, which describe the decrease of CMIN if a modification that is a deviation from the initial model is introduced; procedure proposed e.g. by Byrne, 2016). According to the high sizes of MIs, we successively introduced correlated errors, first between the items “attention” and “respect” and second between “respect” and “communication”. These error covariances were held equal between the two language groups to allow for comparable models if configural invariance was evaluated. The modifications led to an acceptable model fit due to CFI and RMSEA. This model was used as a baseline from which to proceed to the further steps of evaluating metric and scalar invariance. Restricting factor loadings of indicators to being equal between the language groups significantly decreased model fit according to the change in all goodness of fit statistics, so that metric invariance was rejected. Due to its reasonable MI (greater than 3.84, e.g., Kelloway, 2015), the loading of “autonomy” differed between the languages (Table 4). Restricting indicators’ intercepts to being equal between the language groups again significantly decreased model fit according to the change of all goodness of fit statistics. Thus, scalar invariance was not supported. Modification indexes were significant (greater than 3.84, e.g., Kelloway, 2015) for four of seven thresholds (Table 5). The BIC values increased accordingly (Table 6) when restrictions were introduced, which supports the results obtained by the change of other fit indexes.

**Figure 2.**
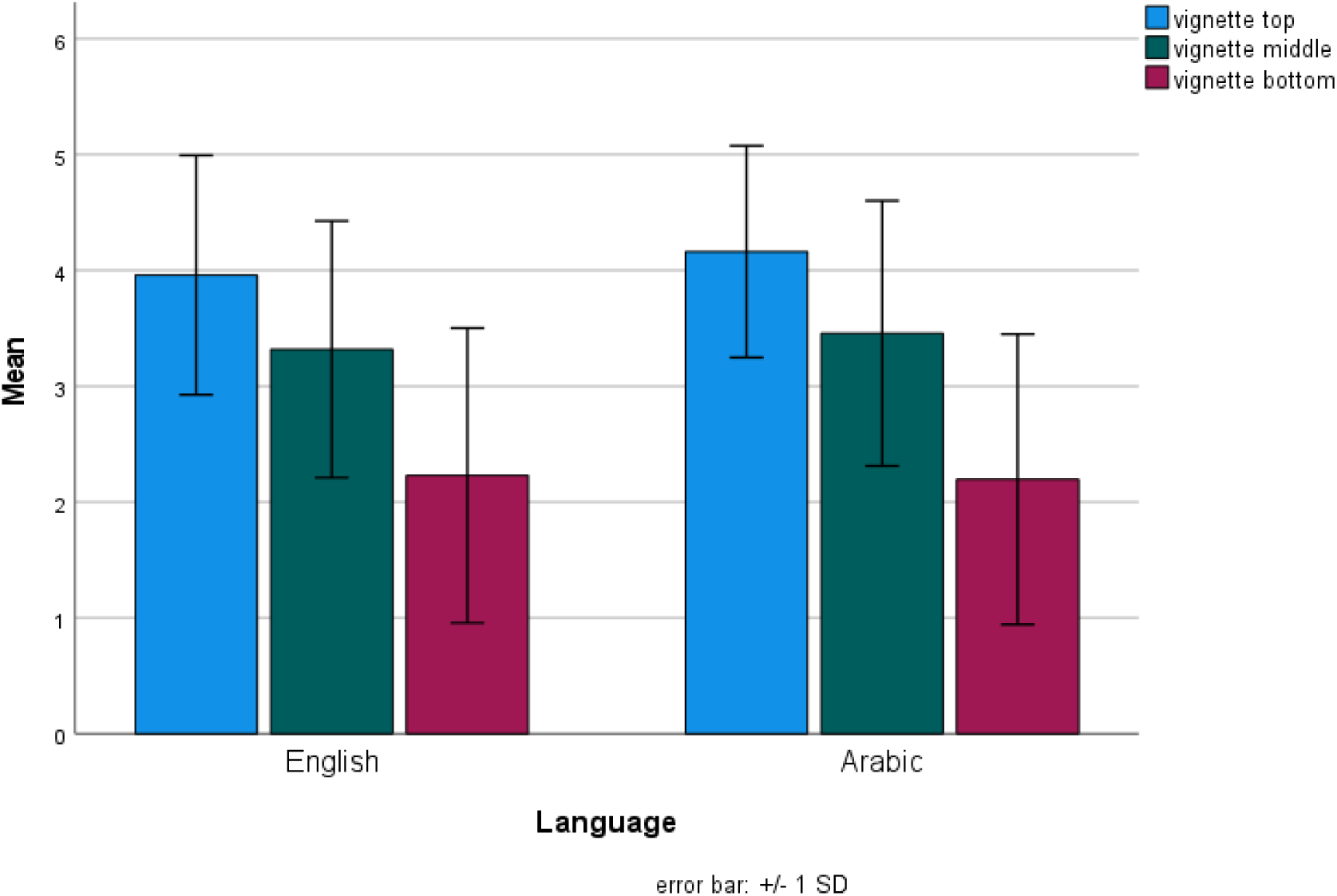
Evaluation of the Top, Middle and Bottom Vignettes by Language

**Figure 3.**
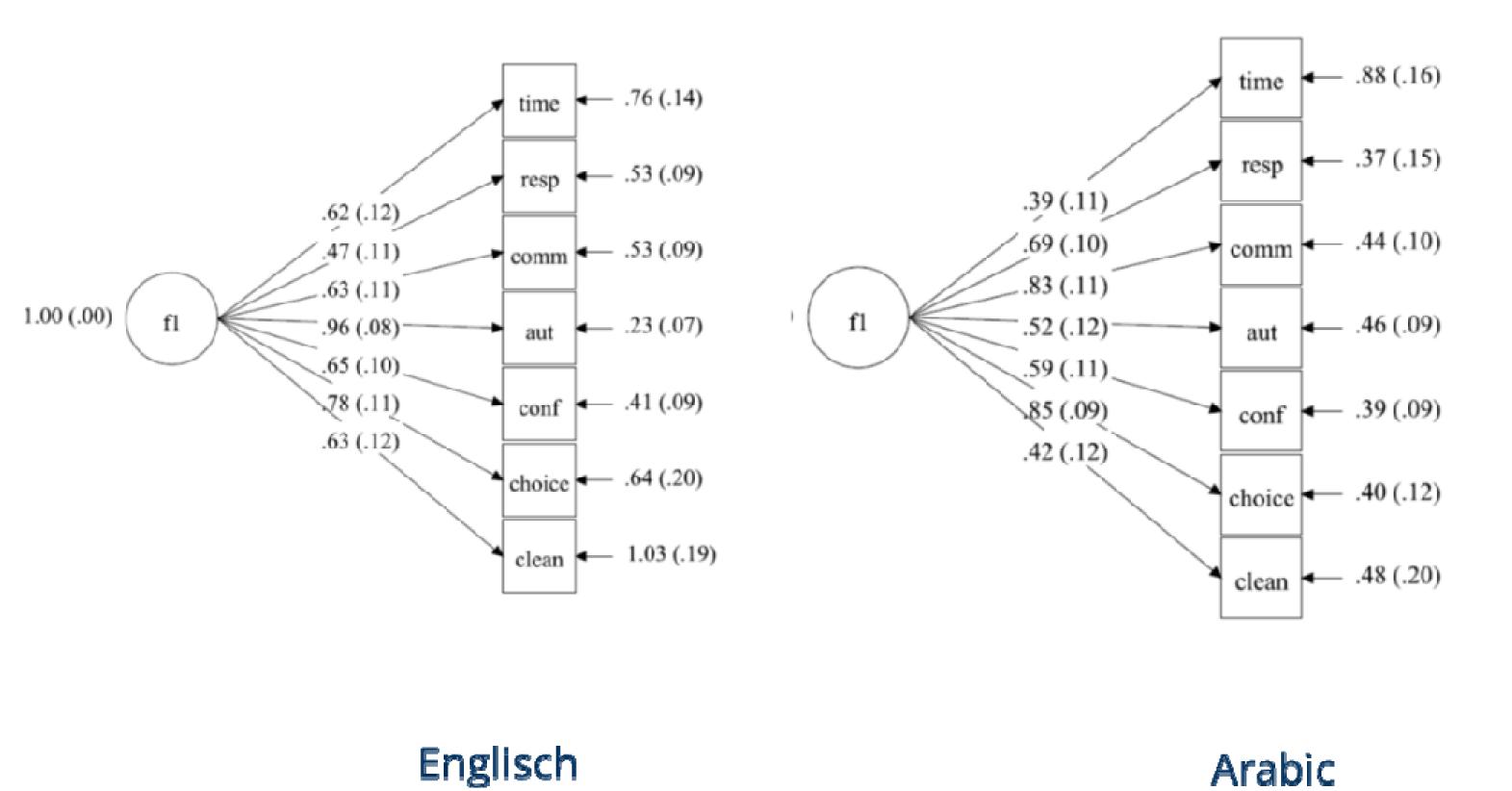
Initial HSR MGCFA Configural Model in English and Arabic Languages Note. f1 Factor HSR

**Table 4.**
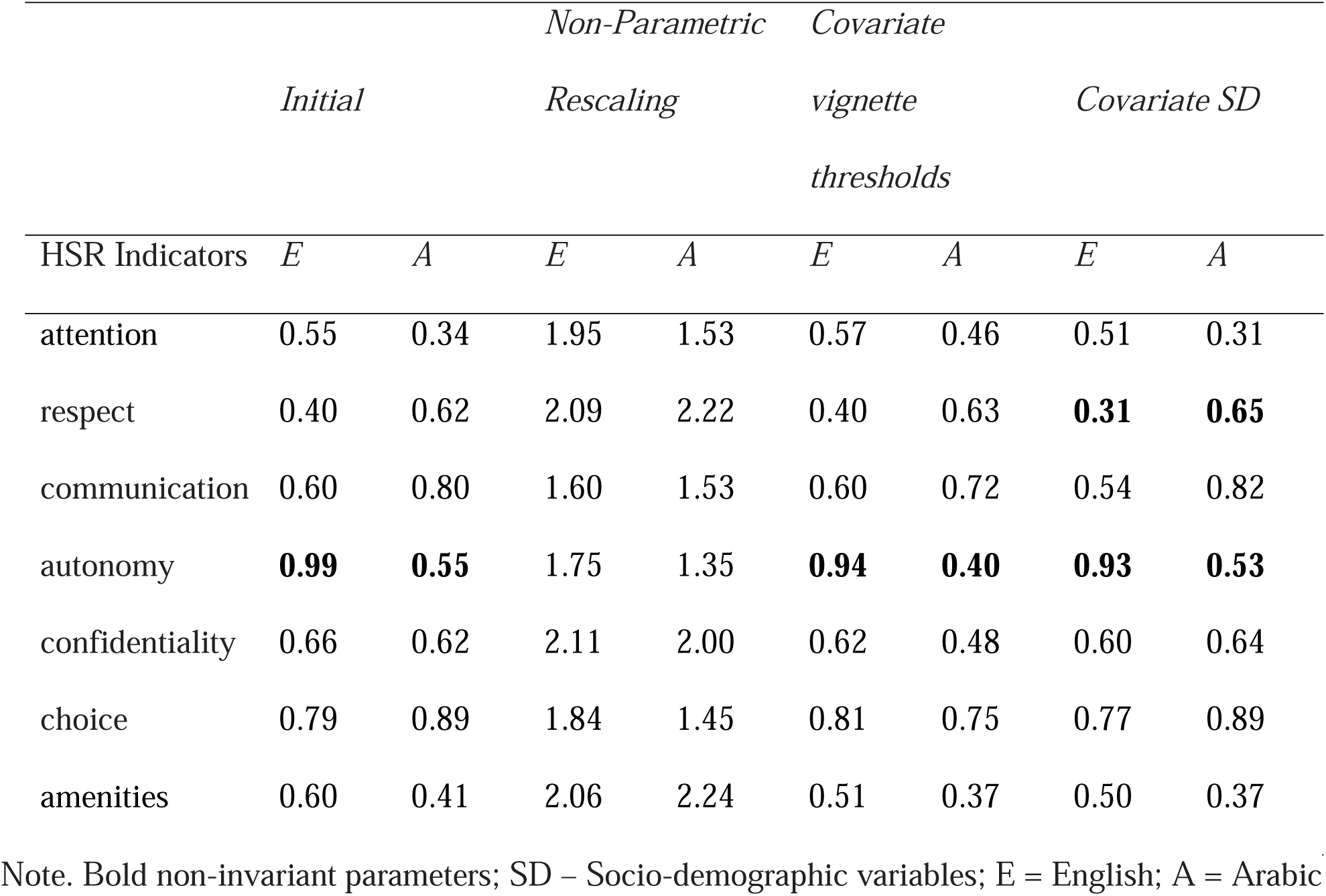
Non-Standardized Loadings of Configural Models of HSR and Covariate Models

**Table 5.** Non-Standardized Intercepts of Configural Models of HSR and Covariate Models

| HSR Indicators | *Initial* |  | *Non-Parametric Rescaling* |  | *Covariate vignette thresholds* |  | *Covariate SD* |  |
| --- | --- | --- | --- | --- | --- | --- | --- | --- |
|  | *E* | *A* | *E* | *A* | *E* | *A* | *E* | *A* |
| attention | **3.64** | **3.46** | 2.80 | 2.23 | 2.81 | 4.00 | 3.68 | 3.34 |
| respect | 4.22 | 4.28 | 3.71 | 3.32 | 2.62 | 2.78 | 4.29 | 4.22 |
| communication | **4.00** | **3.83** | 3.19 | 2.46 | **4.32** | **2.96** | 3.87 | 3.65 |
| autonomy | 3.78 | 3.79 | 2.94 | 2.45 | 3.84 | 3.89 | 3.63 | 3.68 |
| confidentiality | **3.97** | **4.21** | 3.07 | 3.03 | 2.70 | 3.07 | 3.81 | 4.12 |
| choice | 3.76 | 3.76 | 2.92 | 2.37 | 1.04 | 2.61 | 3.69 | 3.97 |
| amenities | **3.95** | **4.49** | 3.56 | 3.63 | 2.35 | 5.45 | 4.16 | 4.49 |

Note. Bold non-invariant parameters; SD – Socio-demographic variables; E = English; A = Arabic

**Table 6.**
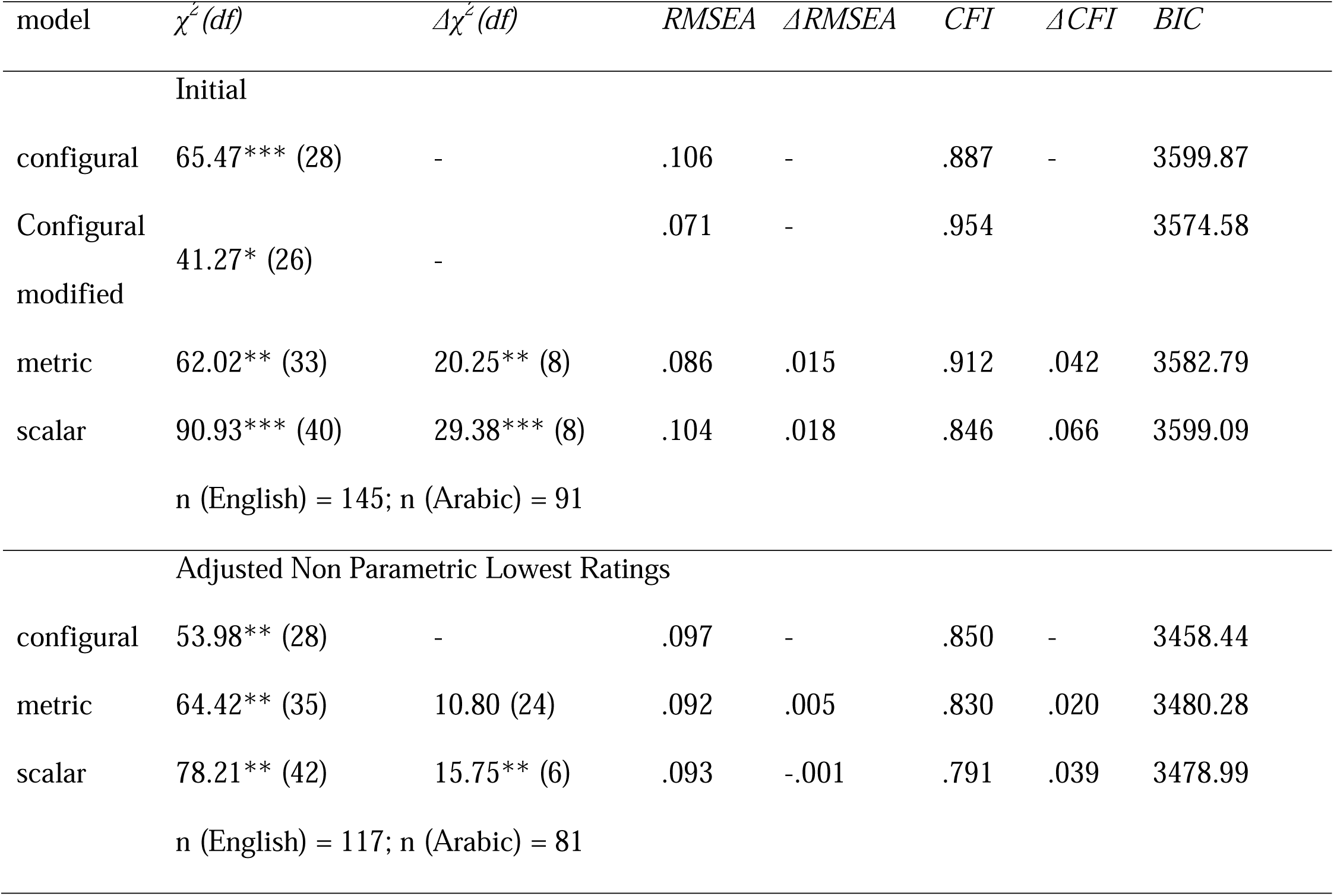
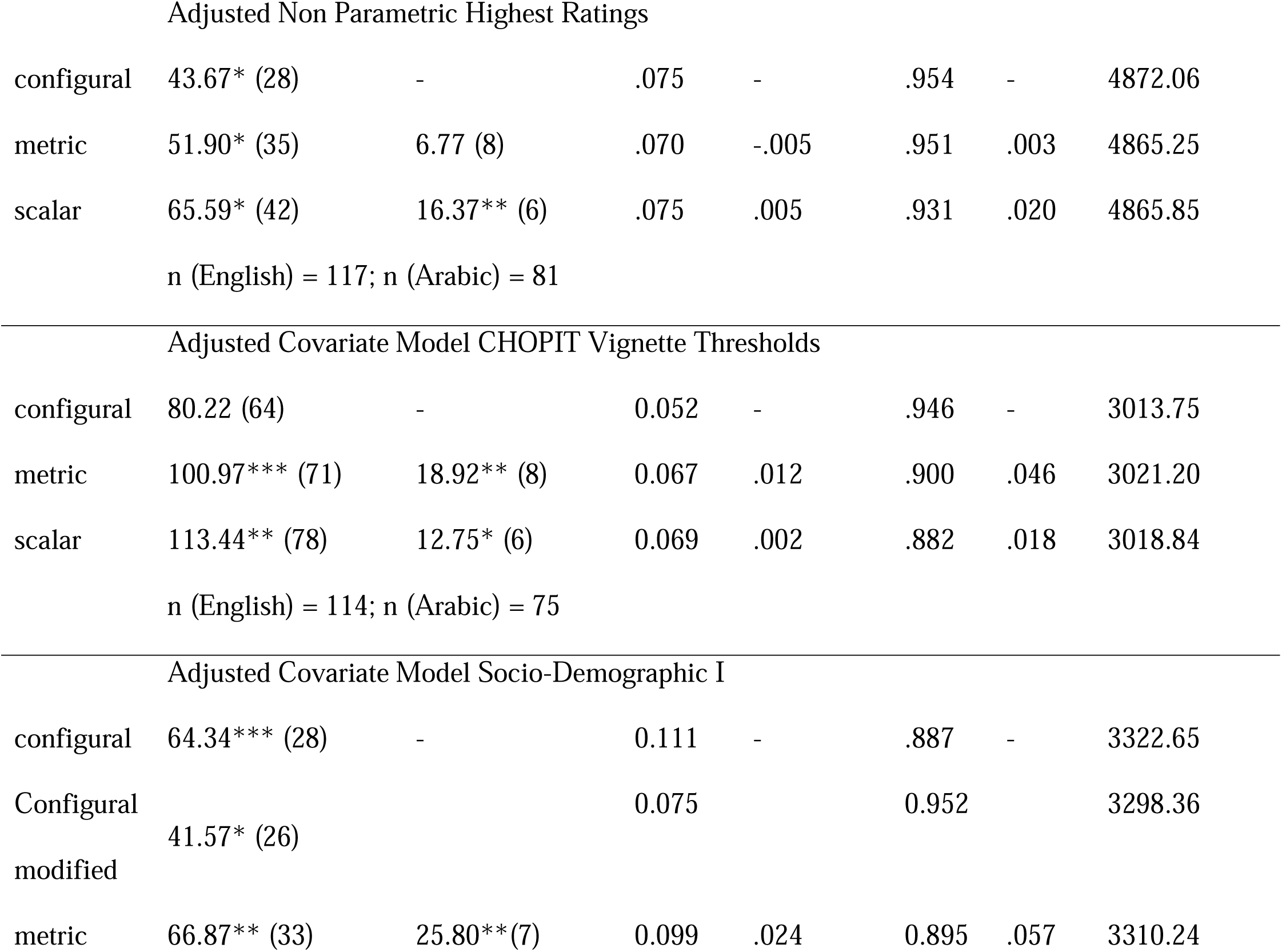
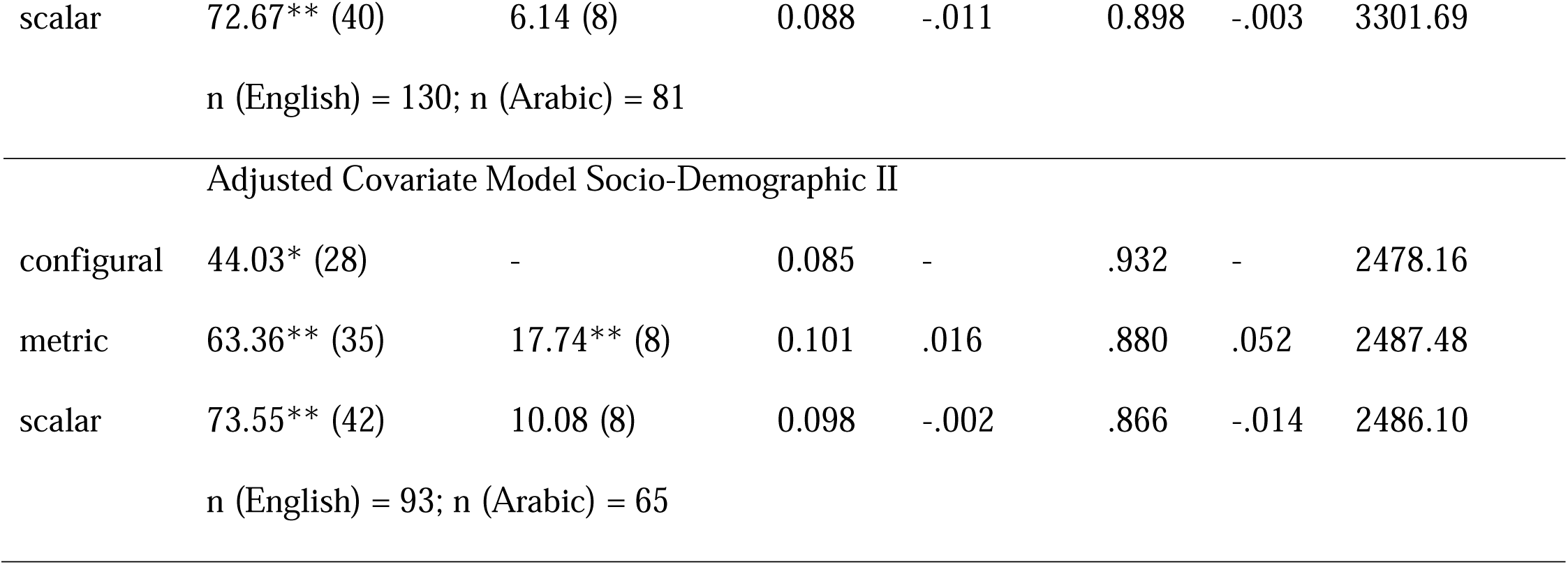
Measurement Invariance Analysis for HSR without and with Adjustment

The results of the analyses with ordinal data are reported in Appendix 5, whereas configural, metric and scalar measurement invariance of the HSR instrument was rejected.

We also conducted a robustness check accounting for sample clustering in the data. Only 18 persons were sharing a room (n = 9 rooms) and nesting in rooms is therefore rather negligible. The clustering effect of reception and accommodation centres on the CFA model for the responsiveness was controlled for by the two-level random intercept CFA analysis (Muthén & Muthén, 2014). The results provide no significant intercept variance on the level of reception and accommodation centres (Var = 0.04, SE = 0.06). To additionally consider clustering when implementing MGCFA, we conducted the analyses using combined weights for the clustered data for rooms and facilities. The results were very similar to those obtained with data not weighted with respect to the model fit and its change (e.g., configural: CMIN (df = 28) = 64; RMSEA = .106; CFI = 0.841; BIC = 3672.13). Therefore, clustering effects did not change the results and we therefore continued the remaining analyses using not weighted data.

Overall, we conclude that the measurement invariance was rejected and therefore violated for the HSR instrument in our sample, whilst HSR did not exhibit configural, metric and scalar measurement invariance.

#### Non-Parametric Rescaling as Basis of Measurement Invariance Analysis

When using rescaled C variables (see Equation 1 and section 2.3) and the lowest possible ratings, measurement invariance could not be improved, as compared with not rescaled data (Table 6). We do not provide further details for this model, i.e. parameters in Tables 4 and 5. However, with the highest possible ratings, acceptable model fit was obtained for configural model according to CFI and RMSEA and there was no significant decrease in model fit statistics in the metric model. With respect to the scalar invariance, the change of CMIN was significant, the change of CFI was of a border value and the change of RMSEA was not significant. With the ordinal data analysis, metric and scalar measurement invariance were supported (Appendix 5). BIC provided no considerable change among all models. This was a better result than with the initial model.

#### MGCFA Covariate Models with Predictions from the Parametric CHOPIT Analysis

As described in the data analysis section, we used vignette threshold values for the HSR indicators predicted by the CHOPIT analysis to evaluate how the parametric approach can be combined with measurement invariance analysis. If a threshold had a significant path (regression coefficient) to one or more manifest variables of HSR, it was included as a covariate variable in the MGCFA model. Significant paths on the HSR indicators were found and implemented in the final model for the quality of amenities vignettes (three threshold values) and for the communication vignettes (two threshold values) (Figure 4, see Mplus source code and output in **Appendix 3**). Interestingly, threshold values from the quality of amenities vignettes correlated with most of the indicators of HSR in English and in Arabic (also with those with different content), except the autonomy indicator. Predicted thresholds from the communication vignettes correlated only with the communication self-assessments.

**Figure 4.**
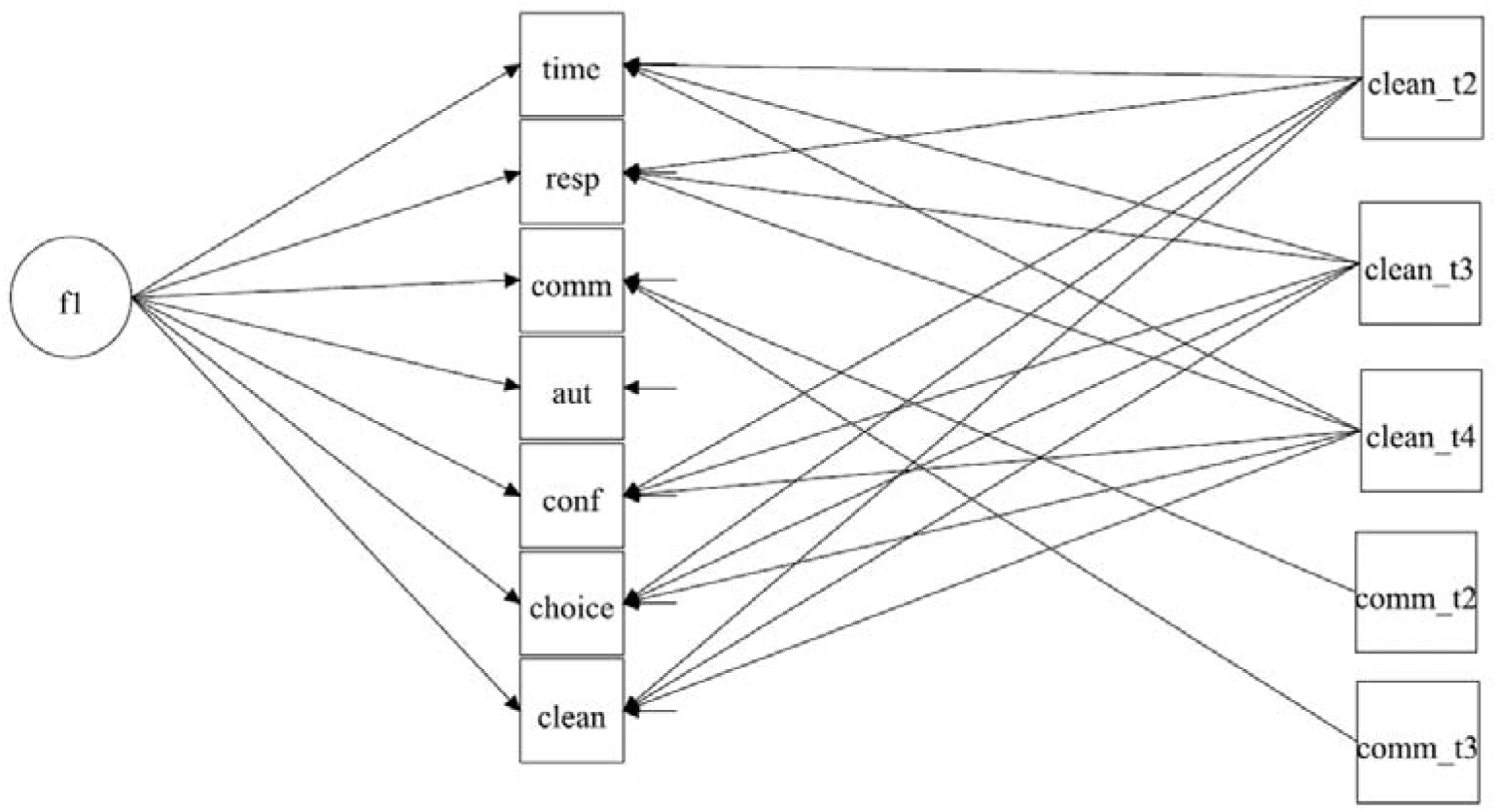
Covariate MGCFA Model of HSR with Threshold Values from the CHOPIT Prediction Note. f1 Factor HSR

**Figure 5.**
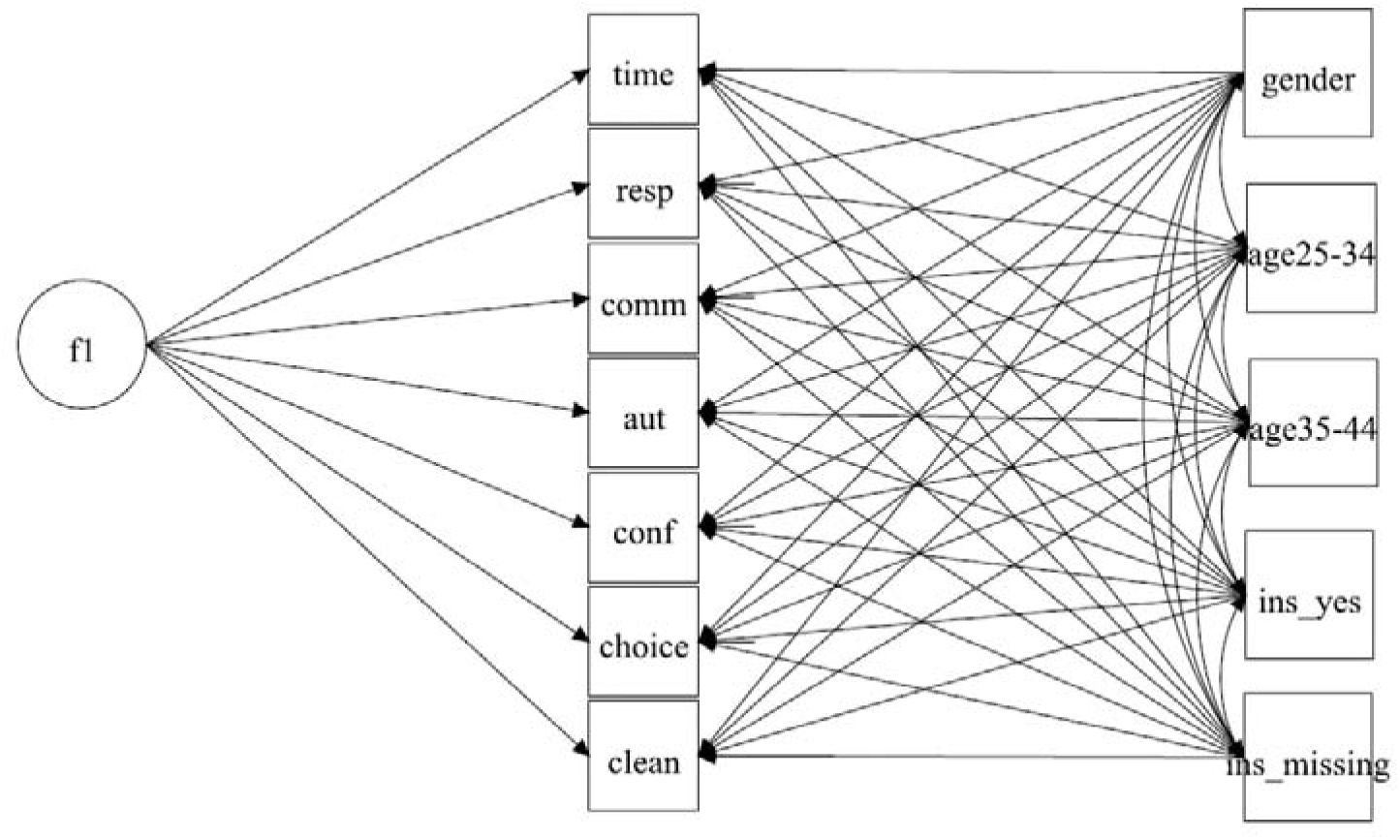
Covariate MGCFA Model of HSR with Socio-Demographic Variables Note. f1 Factor HSR

With the vignette thresholds as covariates, the configural model was associated with a tenable goodness-of-fit, so that configural invariance could be accepted (Table 6, Vignettes CHOPIT Thresholds). Metric invariance had to be rejected, but differences in thresholds as indicators of scalar invariance were reduced similarly to the application of the non-parametric approach. With ordinal analysis, metric and scalar measurement invariance were supported by the BIC statistic (Appendix 5). This means that including information on vignette evaluation in the models when utilizing the parametric approach allowed the acceptance of configural, metric and scalar invariance.

In response to the research questions, we state that with a non-parametric approach, configural, metric and scalar invariance were accepted, particularly if using ordinal models. Therefore, with the adjustments when using anchoring vignettes, we obtained satisfactory and strongly improved results from measurement invariance analysis as compared with the initial HSR model.

#### MGCFA Covariate Models with Socio-Demographic Variables

In the last step, we included socio-demographic variables as covariates in the MGCFA analysis. When education was taken into account this markedly reduced sample size due to missing data and we conducted a separate analysis when education and gender were included as covariate variables.

First, gender, age and possession of a health insurance card were regressed on each of the manifest variables of responsiveness (Figure 4). This model did not provide a tenable model fit (Table 6, Socio Demographic I), so that the configural invariance was rejected. We continued by implementing modifications, as in the case of the initial analysis for HSR. With the same correlated error terms, acceptable model fit was obtained. As in the case of the HSR model, metric invariance was rejected due to the significant change of CMIN and CFI. However, scalar invariance was supported, as restricting intercepts to being equal did not significantly alter the model fit. When looking at the differences of factor loadings according to the MIs (Table 4), the loading of the respect indicators also significantly differed between the languages, but none of the intercepts’ differences contributed to the misfit. Ordinal analyses (Appendix 5) provided comparable results.

Second, we included gender and education as covariate variables in the last MCGFA model (Table 6, Socio Demographic II). The configural model obtained a just acceptable model fit; scalar invariance, but not metric invariance, was supported.

BIC changed in both analyses for socio-demographic variables similarly to CMIN and CFI, showing that the comparisons were not affected by a differing modelling strategy or by sample sizes (like other analyses presented above).

To respond to the research questions, the use of socio-demographic information consistently improved goodness of fit when evaluating scalar invariance, but not when evaluating configural and metric invariance. Since configural and metric invariance should be supported to evaluate scalar invariance, socio-demographic covariates could not be used as mean of control of comparability bias.

## 4. Discussion

We addressed the research question on how to reduce cross-cultural comparability bias in the data on HSR for Arabic- and English-speaking groups of refugees in a federal state in Germany. The cross-cultural comparability bias was evaluated by means of MGCFA measurement invariance analysis including different possibilities of control of RC-DIF. We compared the results of measurement invariance analysis when rescaling data or when including covariates produced with the help of anchoring vignettes. We also compared these possibilities with the inclusion of socio-demographic covariates in the models.

Configural, metric and scalar invariance of HSR between English and Arabic languages was initially violated, which allowed us to test several approaches to influence the non-satisfactory results of measurement invariance analysis. Here, data rescaling based on the non-parametric approach of implementing anchoring vignettes provided the best results and allowed configural, metric – and in ordinal modelling – also scalar measurement invariance to be supported in the models. We also add to previous research (He et al., 2017; Marksteiner et al., 2019) to show that information from anchoring vignettes implemented in the MGCFA models has a strong and positive effect on the results of measurement invariance analysis. Ordinal analyses in particular allowed all levels of measurement invariance to be supported.

We add to previous research by evaluating comparability bias when using information gained from the parametric modelling approach for correction of RC-DIF by means of anchoring vignettes. We implemented a two-step procedure: 1) predict vignette threshold parameters from CHOPIT analysis and 2) introduce them into the MGCFA covariate models.

Besides the use of anchoring vignettes, we included socio-demographic information on gender, age, education and health insurance in the measurement invariance analysis (this information was included in the parametric approach as well). This was not associated with an improved model fit (or bias reduction) with respect to configural and metric invariance. However, the comparability of indicators’ thresholds improved, which outperformed the corresponding analyses when using anchoring vignettes. Hence, metric invariance could not be supported and comparability of loadings even worsened if only socio-demographic variables and no information on anchoring vignettes was used. The results with respect to the configural invariance were mixed for socio-demographic information. Unlike the CHOPIT-Analyses reported in the literature (e.g. Rice et al., 2012), we avoided using too many socio-demographic variables due to the small sample size and the homogeneity of our sample with respect to economic factors and living conditions.

We conclude that RC-DIF was present in our data. Rescaling data or including covariates on the basis of anchoring vignettes could improve the cross-cultural comparability of our data, which supports the findings of previous studies (King et al., 2004; Rice et al., 2012, Mõttus et al., 2012; He et al., 2017; Marksteiner et al., 2019). The results also show that RC-DIF as assessed by anchoring vignettes is independent from the effect of socio-demographics variables on data comparability. We can therefore conclude that differences in cognitive response processes at the stage of response when using rating scales (Tourangeau et al., 2000) account for a substantial bias associated with the rejection of configural, metric and scalar measurement invariance.

In addition, the heterogeneity of our refugee samples was associated with the comparability bias: consideration of certain socio-demographic information decreased incomparability of thresholds in indicators. This also supports the assumption that scalar invariance can be due to systematic measurement error stemming from response styles (Kline, 2016; Cheung & Rensvold, 2002). It is also important to bear in mind that scalar invariance is the last step in the measurement invariance analysis and relies on configural and metric invariance. As the latter were both violated when socio-demographic information was used, these cannot be used as a standalone method for bias control. Further research can consider other and more socio-demographic variables when large sample sizes are used in order to investigate the possibility of their use when evaluating measurement invariance.

The improvement in measurement invariance results was obtained for non-parametric and parametric approaches to implement vignette data in self-evaluation data, although we were not able to implement the full set on vignettes for every indicator. The non-parametric approach used data on a selection of vignettes (bottom, middle and top) for every sampled person. The parametric approach was based on predicted full vignette information from only one respondents’ subgroup and not from the entire sample. However, for the parametric approach, we included information on vignettes from two indicators only (amenities and communication), because the vignette evaluations for other indicators did not correlate with any other self-evaluations. This might point to limited vignette consistency, as response patterns for vignettes and self-evaluation were different. Vignette consistency was given for vignette indicators we included into the modeling. The vignettes on indicator “amenities” not only exhibited consistency with the corresponding self-evaluations, but also with the self-evaluations of other HSR indicators. Therefore, for the control of RC-DIF in measurement invariance analysis, universal anchoring vignettes on topics other than self-assessments would work. Although we conducted analyses that do not rely on the assumption of vignette consistency and vignette equivalence, information gained from anchoring vignettes was useful in increasing model fit for measurement invariance analysis.

In our study, we used data from a unique population-based refugee sample collected in the third largest German federal state. We were able to replicate previous findings by He et al. (2017) for refugee sample and also considered the non-parametric approach. This was possible even though our sample was more heterogeneous than PISA samples used in previous research. The possibility of improving measurement invariance analysis results would be due to the high reliability of HSR in our data, obtained through a careful translation and cognitive pretesting of the instrument. This high reliability also allowed efficient MGCFA analyses despite small sample sizes (Wolf et al., 2013). However, to analyse the potential impact of education on measurement invariance and to implement the information in RC-DIF gained from the parametric approach more productively, replicating research with large samples should be conducted.

Finally, we only investigated exact measurement invariance analysis (Meredith, 1993; Millsap, 2011), although less restrictive methods, such as alignment and a Bayesian approach are available (Asparouchov & Muthén, 2014). We did not use these methods due to their unresolved limitations. The alignment method is suitable in the case of large violations of measurement invariance for single items (Asparouhov & Muthén, 2014) and has been found to be less sensitive in identifying non-comparability problems (Meitinger, 2017). For the Bayesian method, prior information on invariance should be available (Muthén & Asparouhov, 2013), which was not the case in our research.

## 5. Conclusions

Our study contributes to existing research on the comparability of health-related data and the methodology of measurement invariance analysis in several ways. We demonstrate that, in the context of studying a health concept, the implementation of anchoring vignettes can improve the comparability of statistical data in heterogeneous refugee populations. We further provide results that explain RC-DIF as a result of differences in the response process between individuals that use different languages. The adjustments for RC-DIF can therefore improve the results of measurement invariance analysis, which provides a solution to the problems of cross-cultural comparability in survey research (van de Schoot et al., 2015; Meitinger et al., 2020). Use of full sets of anchoring vignettes is also associated with a higher burden on respondents, a longer survey time and increased research costs. Our experiences point to the possibilities for a more economic use of anchoring vignettes. This should be the focus of further research.

## Data Availability

All data produced in the present study are available upon reasonable request to the authors

## 6. Declarations

### Ethics approval and consent to participate

This study was approved by the Ethics Committee of the Medical Faculty of Heidelberg University, Germany (S-516/2017). Informed consent was obtained from all study participants.

### Consent for publication

Not applicable. The manuscript does not contain any personal material or photos of participants

### Availability of data and material

The dataset used during the current study is available from the corresponding author on reasonable request.

### Competing interests

The authors declare that they have no competing interests.

### Funding

This research was supported by the Federal Ministry of Education and Research (01GY1611) and German Research Foundation (ME 3538/10-1; BO 5233/1-1).

### Authors’ contributions

Natalja Menold made substantial contributions to funding acquisition, the conception and design of the work, theory and overview of preliminary research on anchoring vignettes and measurement invariance, conducted data analyses (MANCOVA, SEM, MG-CFA), provided and drafted results and their interpretation, as well as drafted and edited the paper.

Louise Biddle made substantial contributions to the conception and design of the work, theory and preliminary research on Health System Responsiveness and anchoring vignettes, conducted data collection, supported interpretation of results, drafted the sections 2.1 and 2.2 and edited the paper.

Hagen von Hermanni made substantial contributions to the theory and research overview on anchoring vignettes, conducted CHOPIT analysis (glamm), provided and drafted corresponding results, supported interpretation and discussion, and provided Supplementary Material.

Jasmin Kadel made substantial contributions to the theory and research overview on vignettes, conducted CHOPIT analysis (glamm), provided corresponding results, and supported interpretation and discussion.

Kayvan Bozorgmehr made substantial contributions to funding acquisition, the conception and design of the work, conducted data collection, supported drafting the paper and interpretation of results.

All authors reviewed the manuscript.

## Acknowledgements

Not applicable

## 7. List of abbreviations

BIC: Bayesian Information Criterion
CFI: Comparative Fit Index
CHOPIT: Hierarchical Ordered Regression Model
CMIN: chi-square test
DIF: Differential Item Functioning
ESS: European Social Survey
HSR: Health System Responsiveness
ISSP: International Social Survey Program
LVM: Latent Variable Modelling
MANCOVA: Multivariate Analysis of Covariance
MGCFA: Multi-Group Confirmatory Factor Analysis
MI: Modification Indexes
MLR: Robust Maximum Likelihood Estimator
PISA: Programme for International Student Assessment
RC-DIF: Response-Category Differential Item Functioning
RMSEA: Root-Mean-Square Error of Approximation
SAGE: Global Ageing and Adult Health Survey
SEM: Structural Equation Modelling
SHARE: Survey of Health, Ageing and Retirement in Europe
TIMMS: Trends in International Mathematics and Science Study
WLS: Wisconsin longitudinal Study
WHO: World Health Organization
WHS: World Health Survey

## Appendix 1

### Predicting C Variables (Non-Parametric Approach)

See Wand, J., & King, G. (2007). Anchoring vignetttes in R: A (different kind of) vignette. Retrieved from http://wand.stanford.edu/anchors/doc/anchors.pdf

\# s_att to s_amn: Indicators of HSR; vg: top vignette, vm: medium vignetre, vs buttom vignette

atta <- anchors(s_att ∼ vg+vm+vs, Data, method=“C”)

respa <- anchors(s_res ∼ vg+vm+vs, Data, method=“C”)

coma <- anchors(s_com ∼ vg+vm+vs, Data, method=“C”)

auta <- anchors(s_aut ∼ vg+vm+vs, Data, method=“C”)

cona <- anchors(s_con ∼ vg+vm+vs, Data, method=“C”)

choa <- anchors(s_cho ∼ vg+vm+vs, Data, method=“C”)

amna <- anchors(s_amn ∼ vg+vm+vs, Data, method=“C”)

\## Saving adjusted values (insert + write.sav)

att <- insert(Data, atta, overwrite = TRUE)

resp <- insert(Data, respa, overwrite = TRUE)

com <- insert(Data, coma, overwrite = TRUE)

aut <- insert(Data, auta, overwrite = TRUE)

con<- insert(Data, cona, overwrite = TRUE)

cho<- insert(Data, choa, overwrite = TRUE)

amn<- insert(Data, amna, overwrite = TRUE)

\## Saving Data to SPSS files

library (haven)

write_sav(time, “ Respond_time.sav”)

write_sav(resp, “ Respond_resp.sav”)

write_sav(com, “ Respond_com.sav”)

write_sav(aut, “ Respond_aut.sav”)

write_sav(con, “ Respond_con.sav”)

write_sav(cho, “ Respond_cho.sav”)

write_sav(amn, “ Respond_amn.sav”)

### Analysis Vignette Ordering

voengl<-anchors.order(∼vs+vm+vg, Data_engl)

summary(voengl,top=10,digits=3)

voarb<-anchors.order(∼vs+vm+vg, Data arabic)

summary(voarb,top=10,digits=3)

## Appendix 2

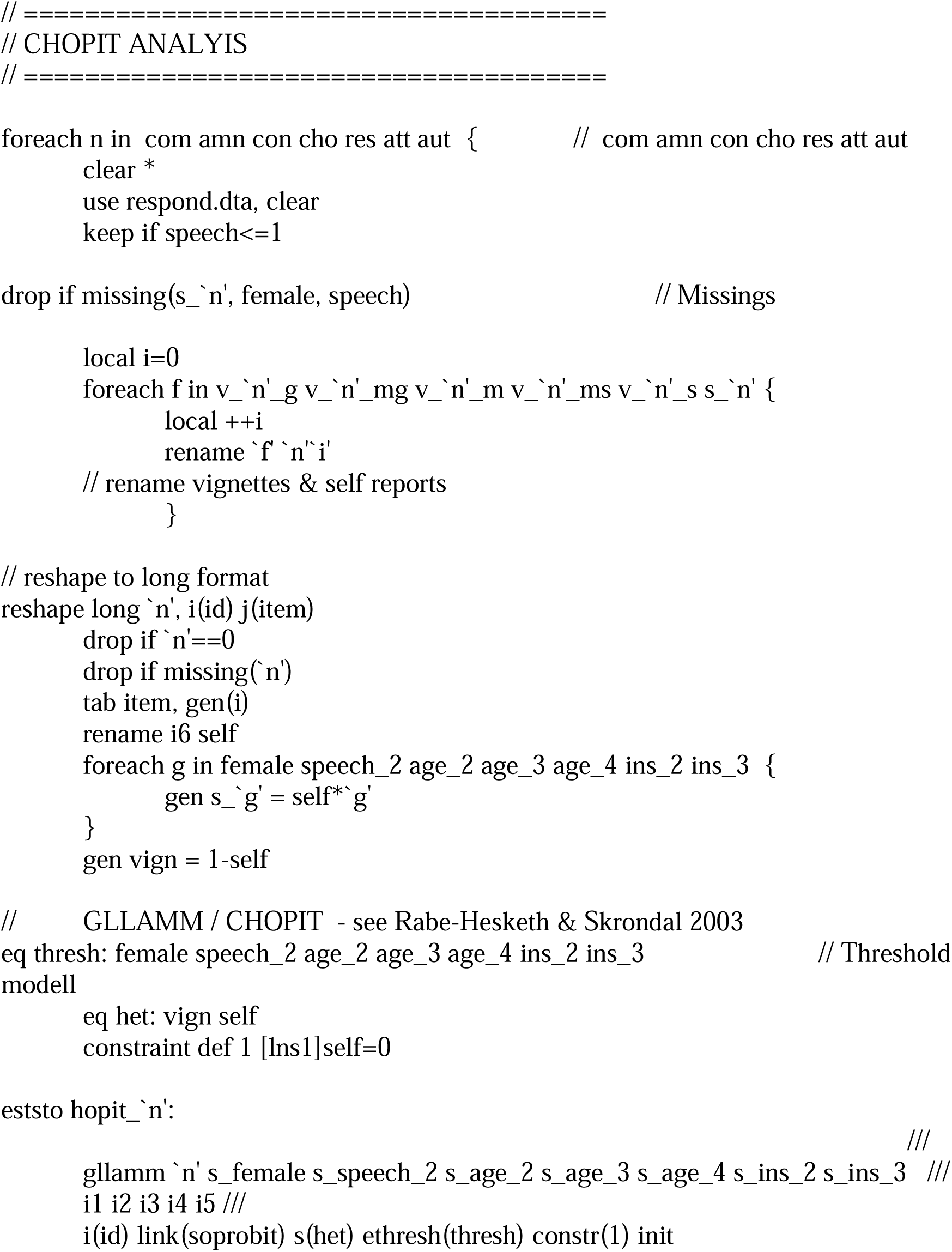
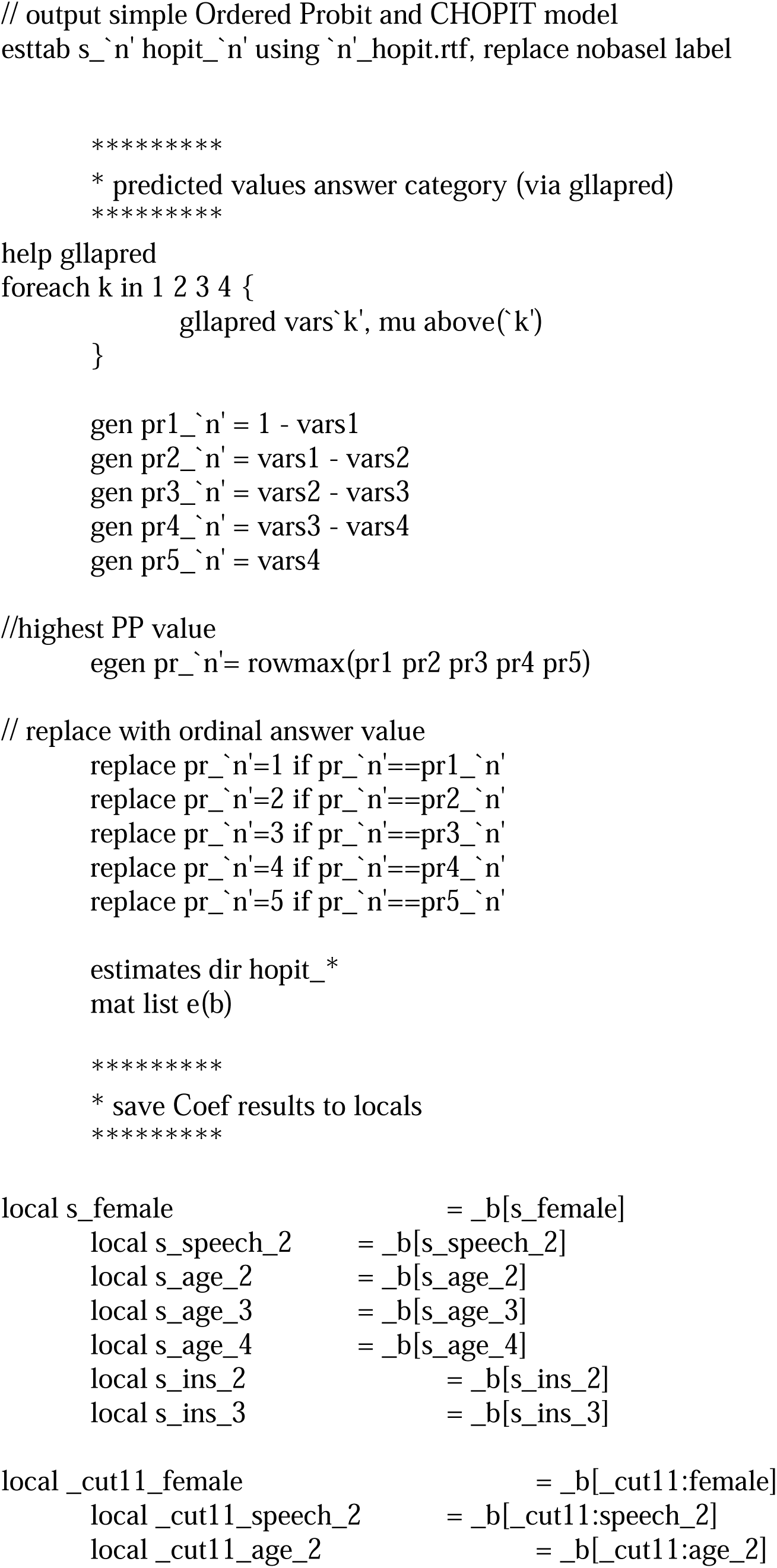
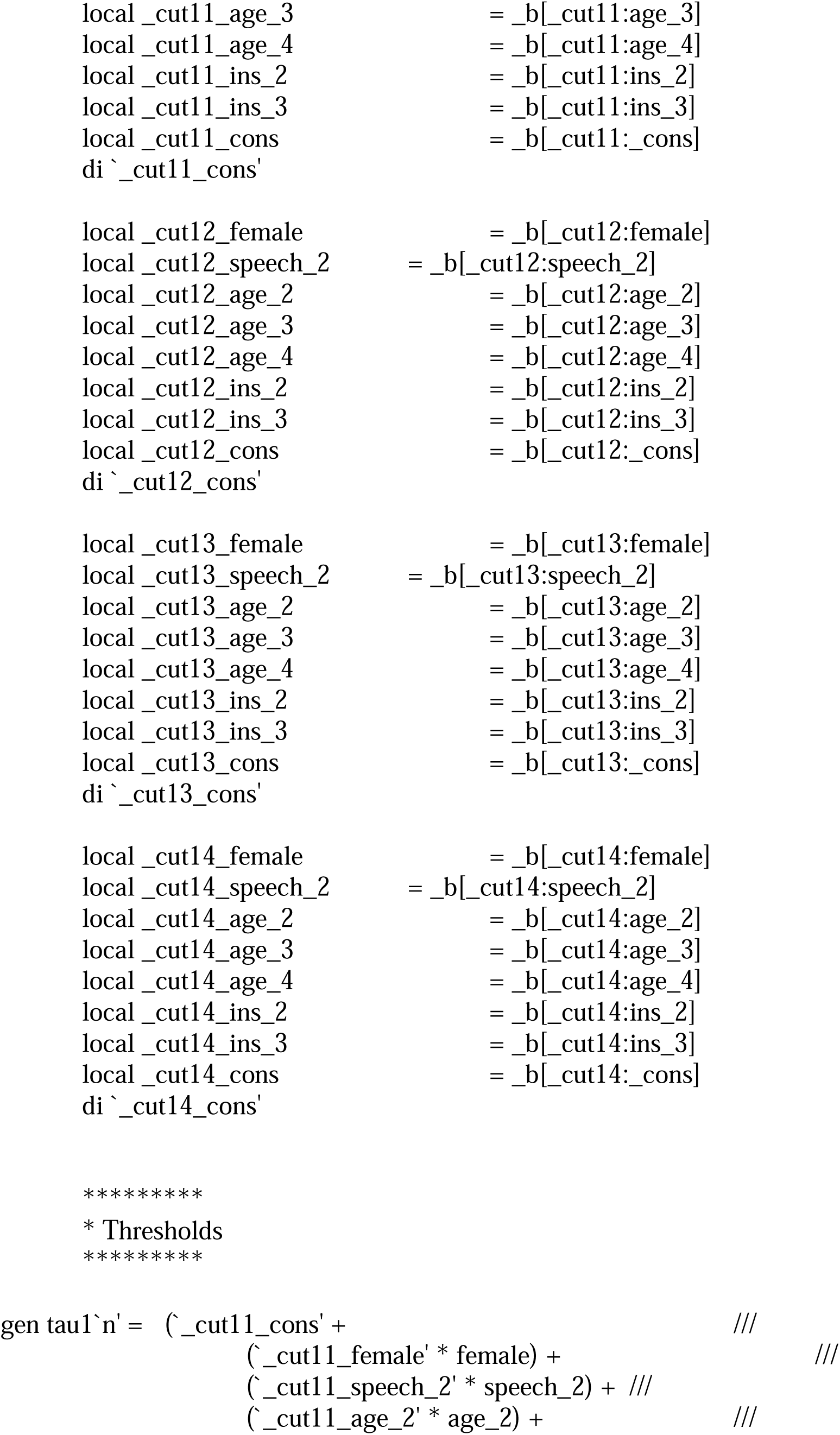
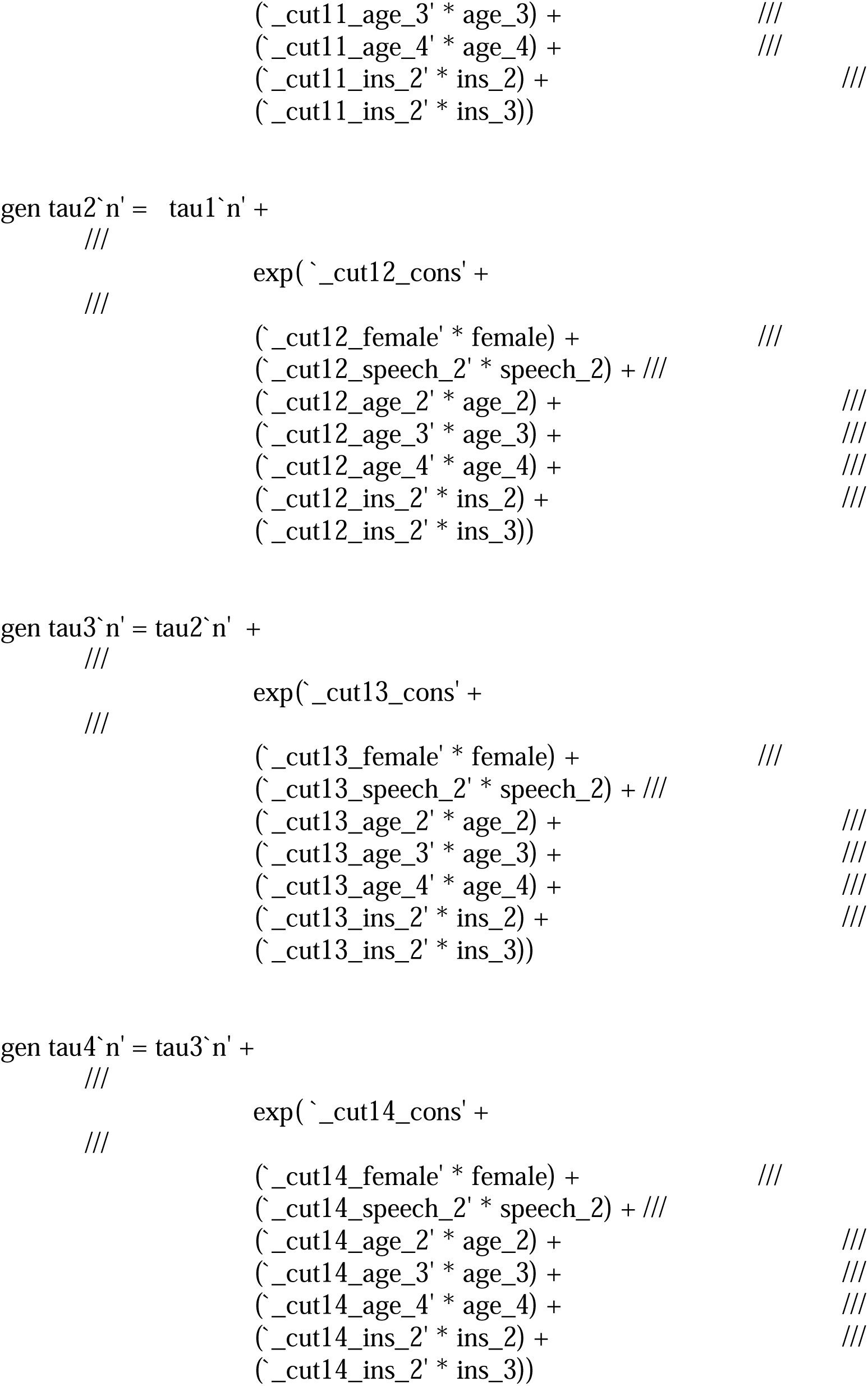
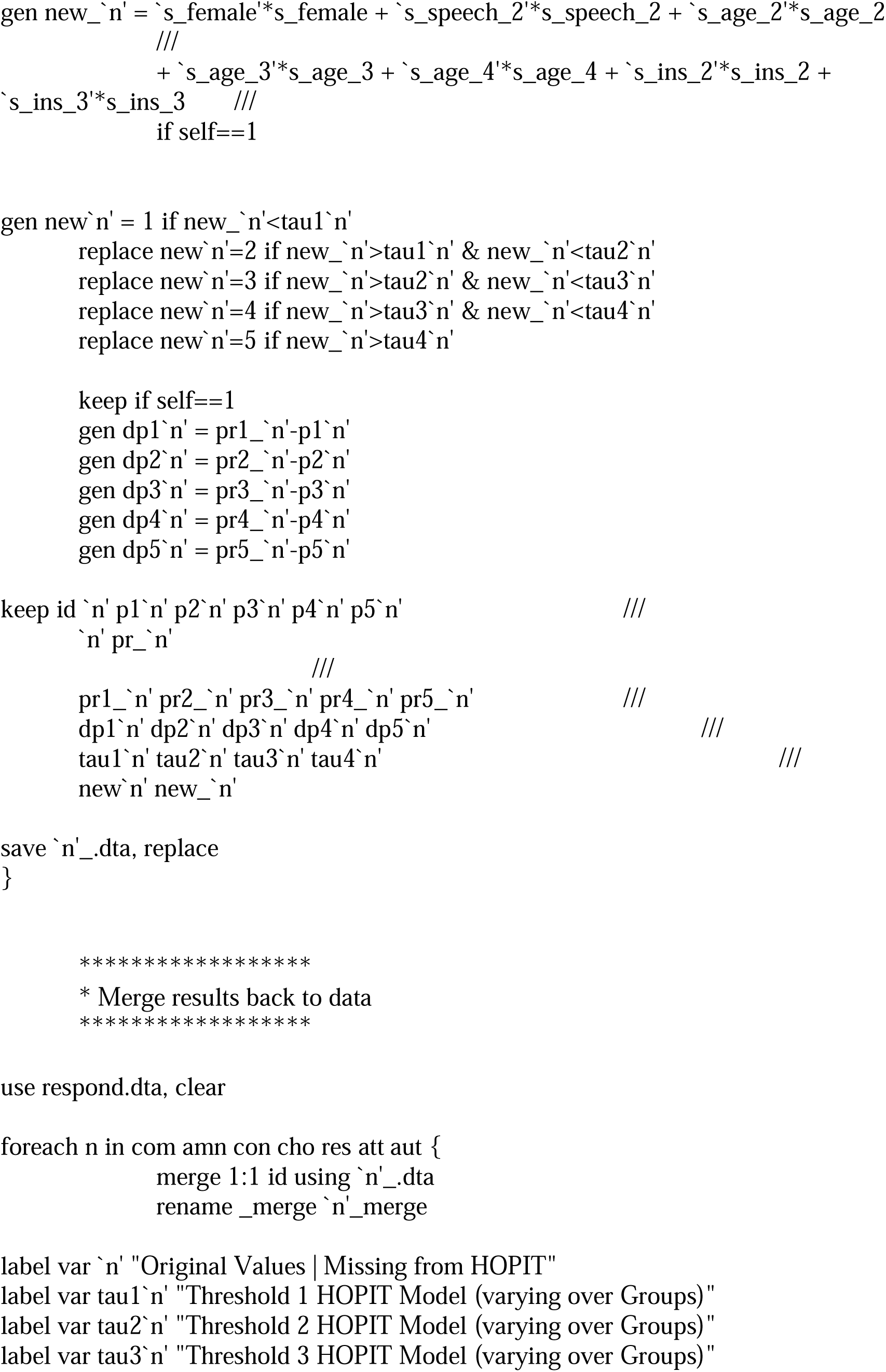
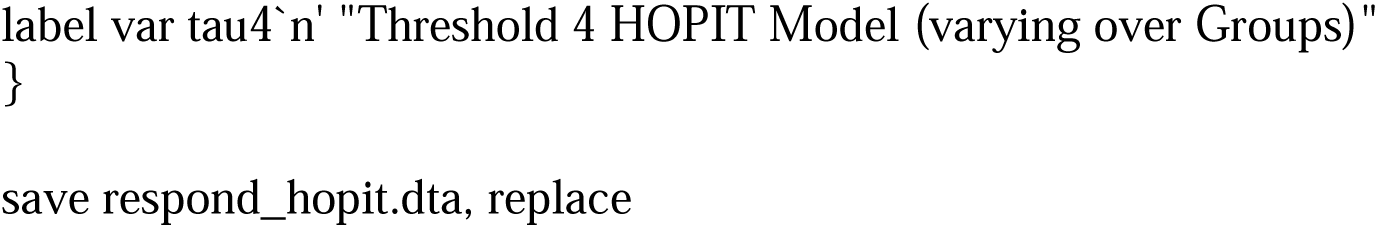

## Appendix 3

Mplus Source Data for Covariate Model CHOPIT Thresholds

TITLE: Model_resp. !Title of the model

DATA: FILE = Resp.dat; !Where is the data

VARIABLE: NAMES ARE a1 - a106; !Variables in the data set

GROUPING IS a30(0=0 1=1); ! Grouping variable: 0 = English, 1 = Arabic

USEVARIABLES a2-a8 a81-a83 a77 a78; !Variables used in the model; a2-a8: HSR indicators; a81-a83 amenities vignettes thresholds; a 77 a78 communication vignette thresholds, obtained by GLAMM analysis, see Appendix 2

MISSING ALL (-99); ! Declaration of missing values

ANALYSIS:

ESTIMATOR = MLR;

MODEL:

F1 BY a2* a3-a8; !Measurement part: CFA of HSR

a2-a3 a6-a8 ON a81-a83;

a4 ON a77 a78; !Structural part: HSR indicators regressed on threshold values

F1@1; !Factor variance set to 1;

[F1@0]; !Factor mean set to 1; necessary to compare intercepts

Model 1: !Specification for the Model Arabic, Factor loadings and intercepts are

!different

F1 BY a2* a3-a8;

[a2-a8];

OUTPUT: STANDARDIZED (STDYX);

MODINDICES (3.84);

## Appendix 4

Mplus Source Data for Covariate Model Socio-Demographic Variables

TITLE: Model_resp. !Title of the model

DATA: FILE = RespSD.dat; !Where is the data

VARIABLE: NAMES ARE a1 - a128; !Variables in the data set

GROUPING IS a30(0=0 1=1); ! Grouping variable: 0 = English, 1 = Arabic

USEVARIABLES a2-a8 a35 a122 a123 a127 a128; !a2-a8 HSR indicators; a35 gender; a123 age 25-34 years; a123 age 35-44 years; a127 insurance yeas; a128 insurance missing

MISSING ALL (-99);

ANALYSIS:

ESTIMATOR = MLR;

MODEL:

F1 BY a2* a3-a8; !Measurement part: CFA of HSR

a2-a8 ON a35 a122 a123 a127 a128; !Structural part indicators of HSR are regressed on socio-demographic variables

F1@1;

[F1@0];

a3 with a2 (1); !correlated error terms as model modification

a3 with a4 (2); !correlated error term as model modification

Model 1:

F1 BY a2* a3-a8;

F1@1;

[a2-a8];

OUTPUT: STANDARDIZED (STDYX);

MODINDICES (3.84);

## Appendix 5

### Measurement Invariance Analyses for Ordinal Data

We conducted different analyses with specifying data as ordered categorical variables. First, we used the default for measurement invariance analysis of Mplus software with the specification of measurement invariance analysis in the ANALYSIS command (Muthén & Muthén, 2014). The configural model obtained a poor model fit according to CMIN and RMSEA and had therefore to be rejected (Table 1). Metric invariance model could not be evaluated due estimation problems. Alternatively, measurement invariance analysis for categorical data can be conducted by means of mixture modelling, which we implemented. For these analyses, only BIC statistics are available for the evaluation of model fit. Mixture modelling revealed similar results to the analyses with MLR reported in the main text (Table 6 in main text) and metric and scalar invariance of the initial ordinal model had to be rejected. When using rescaling data from anchoring vignettes, configural, metric and scalar invariance could not be rejected. Hence, RMSEA for the configural was more tenable in the non-categorical analysis (Table 6, main text).

This default analysis of Mplus for measurement invariance analysis did not allow for covariates to be introduced into the models. The covariate models were evaluated using mixture modelling as well. Configural measurement invariance is improved in the models with predicted vignette thresholds and socio-demographic variables gender, age, and medical insurance card (Socio-Demographic I). According to the change in the BIC statistic, metric and scalar invariance hold in the model with CHOPT predictions. For socio-demographic covariates, the modelling results are strongly comparable to those obtained with the analyses reported in the main text.

We can therefore conclude that using anchoring vignettes produces even more satisfactory results with respect to the support of metric and scalar measurement invariance of HSR indicators. However, these analyses have the disadvantage of a limited number of statistics for comparison purposes. In addition, configural measurement invariance cannot be evaluated when using BIC.

**Table 1.** Ordinal Measurement Invariance Analysis for HSR and Non-Parametric Adjustment

| model | $\chi^2(df)$ | $\Delta\chi^2(df)$ | *RMSEA* | $\Delta RMSEA$ | *CFI* | $\Delta CFI$ |
| --- | --- | --- | --- | --- | --- | --- |
| Initial |  |  |  |  |  |  |
| configural | 117.83*** (28) | - | 0.165 | - | .947 | - |
| metric | NA (estimation problems) | - |  |  |  |  |
| scalar | 178.64*** (54) | 75.74*** (26) | 0.140 | - | .927 | - |
| Non parametric highest ratings |  |  |  |  |  |  |
| configural | 50.79** (28) | - | .091 | - | .993 | - |
| metric | 60.52** (34) | 10.30 (6) | .089 | .003 | .992 | .001 |
| scalar | 71.14** (54) | 12.20 (6) | .089 | .000 | .990 | .002 |

Note. MGCFA Model with categorical indicators and MODEL = CONFIGURAL METRIC SCALAR in the ANALYSIS command. This analysis is not available for the models with covariates.

**Table 2.** Sample-Size Adjusted BIC for Measurement Invariance Analyses with Categorical Data from Mixture Modeling

| Models | Initial | Adjusted Covariate<br>CHOPIT Thresholds | Adjusted Covariate<br>Socio-Demographic I |
| --- | --- | --- | --- |
| Configural | 3481.16 | 2897.48 | 3206.92 |
| Metric | 3488.33 | 2902.11 | 3219.59 |
| Scalar | 3498.39 | 2906.14 | 3212.81 |

Note. BIC Differences $\geq 6$ provide evidence for strong differences (Raftery, 1995)

## Footnotes

1 Additional types of measurement invariance exist, such as strict invariance that assumes equality in error variances across groups. We do not include it in our study because metric invariance is sufficient for the comparison of correlations and scalar invariance is sufficient for the comparison of means, which is also the aim of cross-cultural research. This is similar to the approach taken by other researchers; compare e.g. Hox et al., 2015; Kim et al., 2017.

## Notes

### Competing Interest Statement

The authors have declared no competing interest.

### Funding Statement

the Federal Ministry of Education and Research (01GY1611);
German Research Foundation (ME 3538/10-1; BO 5233/1-1).

### Author Declarations

Ethics Committee of the Medical Faculty of Heidelberg University, Germany gave ethical approval for this work.

## References

Asparouhov T, Muthén B. Multiple-Group Factor Analysis Alignment. Structural Equation Modeling: A Multidisciplinary Journal. 2014;21(4):495–508. doi:10.1080/10705511.2014.919210.

Beauducel A, Wittmann W. Simulation Study on Fit Indexes in CFA Based on Data with Slightly Distorted Simple Structure. Struct Equ Modeling. 2005;12:41–75.

Behr D. Translation studies and internationally comparative survey research: quality assurance as object of a process analysis. GESIS-Schriftenreihe Vol. 2. Bonn: GESIS; 2009. https://www.ssoar.info/ssoar/handle/document/26125. Accessed 06 Apr 2022.

Biddle L, Hintermeier M, Mohsenpour A, Sand M, Bozorgmehr K. Monitoring der Gesundheit und Gesundheitsversorgung geflüchteter Menschen in Sammelunterkünften: Ergebnisse des bevölkerungsbezogenen Surveys. J Health Monit. 2021; doi:10.25646/7862.

Biddle L, Menold N, Bentner M, Nöst S, Jahn R, Ziegler S, Bozorgmehr K. Health monitoring among asylum seekers and refugees: a state-wide, cross-sectional, population-based study in Germany. Emerg Themes Epidemiol. 2019; doi:10.1186/s12982-019-0085-2.

Byrne B. Structural Equation Modeling with Mplus: Basic Concepts, Applications, and Programming (Multivariate Applications). London: Taylor & Francis; 2016.

Chen FF. Sensitivity of Goodness of Fit Indexes to Lack of Measurement Invariance. Struct Equ Modeling. 2007;14:464–504.

Cheung GW, Rensvold RB. Evaluating Goodness-of-Fit Indexes for Testing Measurement Invariance. Struct Equ Modeling. 2002;9:233–55.

Davidov E, Meuleman B, Cieciuch J, Schmidt P, Billiet J. Measurement Equivalence in Cross-National Research. Annu Rev Sociol. 2014;40:55–75.

Davidov E, Cieciuch J, Schmidt P. The Cross-Country Measurement Comparability in the Immigration Module of the European Social Survey 2014-15. Surv Res Methods. 2018;12:15–27.

Davidov E, Meuleman B, Billiet J, Schmidt P. Values and Support for Immigration: A Cross-Country Comparison. Eur Sociol Rev. 2008;24:583–99.

D’uva TB, Lindeboom M, O’Donnell O, van Doorslaer E. Education-related inequity in healthcare with heterogeneous reporting of health. J R Stat Soc Ser A Stat Soc. 2011; doi: 10.1111/j.1467-985X.2011.00706.x.

Dong Y, Dumas D. Are personality measures valid for different populations? A systematic review of measurement invariance across cultures, gender, and age. Personality and Individual Differences. 2020;160:109956. doi:10.1016/j.paid.2020.109956.

Greene WH, Harris MN, Knott RJ, Rice N. Specification and testing of hierarchical ordered response models with anchoring vignettes. Journal of the Royal Statistical Society: Series A (Statistics in Society). 2021;184(1):31–64. doi:10.1111/rssa.12612.

Gregorich SE. Do self-report instruments allow meaningful comparisons across diverse population groups? Testing measurement invariance using the confirmatory factor analysis framework. Medical Care. 2006; doi:10.1097/01.mlr.0000245454.12228.8f.

Grol-Prokopczyk H, Freese J, Hauser RM. Using anchoring vignettes to assess group differences in general self-rated health. J Health Soc Behav. 2011;52:246–261.

Hadler P, Neuert C, Lenzner T, Stiegler A, Sarafoglou A, Bous P, Reisepatt N, Menold N. RESPOND – Improving Regional Health System Responses to the Challenges of Migration Through Tailored Interventions for Asylum-Seekers and Refugees: Kognitiver Pretest. GESIS Projektbericht. 2017; doi:10.17173/pretest69.

Harrison S, Henderson J, Alderdice F, Quigley MA. Methods to increase response rates to a population-based maternity survey: a comparison of two pilot studies. BMC Medical Research Methodology. 2019;19(1):65. doi:10.1186/s12874-019-0702-3.

Harkness JA, Villar A, Edwards B. Translation, Adaptation, and Design. In: Harkness, JA, Braun M, Edwards B, Johnson TP, Lyberg L, Mohler PP, Pennell B-E, Smith TW, editors. Survey Methods in Multinational, Multiregional, and Multicultural Contexts. New Jersey: Willey; 2010. p. 117–140.

He J, Buchholz J, Klieme E. Effects of anchoring vignettes on comparability and predictive validity of student self-reports in 64 cultures. J Cross Cult Psychol. 2017;48:319–34.

Holland PW, Wainer H. Differential Item Functioning. Lawrence Erlbaum Associates, Hove and London; 1993.

Hopkins DJ, King G. Improving Anchoring Vignettes: Designing Surveys to Correct Interpersonal Incomparability. Public Opinion Quarterly. 2010;74(2):201–222. http://www.jstor.org/stable/40660640.

Hox JJ, De Leeuw ED, Zijlmans EAO. Measurement Equivalence in Mixed Mode Surveys. Front Psychol. 2015; doi:10.3389/fpsyg.2015.00087.

Hu L, Bentler PM. Cutoff Criteria for Fit Indexes in Covariance Structure Analysis: Conventional Criteria Versus New Alternatives. Structural Equation Modeling: A Multidisciplinary Journal. 1999;6:1–55.

Jöreskog KG. Simultaneous Factor Analysis in Several Populations. Psychometrika. 1971;36:409–26.

Kyllonen, PC, Bertling, JP. Innovative questionnaire assessment methods to increase cross-country comparability. In: Rutkowski L, Von Davier M, Rutkowski D, editors. ASSESSMENT IN EDUCATION: PRINCIPLES, POLICY & PRACTICE 533 Handbook of international large-scale assessment: Background, technical issues, and methods of data analysis (pp. 277–286). Boca Raton, FL: CRC Press; 2014.

Kelloway EK. Using Mplus for structural equation modeling: A researcher’s guide. 2nd edition. SAGE; 2015.

Kim ES, Cao C, Wang Y, Nguyen DT. Measurement Invariance Testing with Many Groups: A Comparison of Five Approaches. Structural Equation Modeling: A Multidisciplinary Journal. 2017;24(4):524–544. doi:10.1080/10705511.2017.1304822.

King G, Wand J. Comparing incomparable survey responses: Evaluating and selecting anchoring vignettes. Polit Anal. 2007;15:46–66.

King G, Murray, CJL, Salomon JA, Tandon, A. Enhancing the validity and cross-cultural comparability of measurement in survey research. Am Polit Sci Rev. 2004;98:191–207.

Kline RB. Principles and Practices of Structural Equation Modeling. New York: The Guilford Press; 2016.

Lee S, Vasquez E, Ryan L, Smith J. Measurement Equivalence of Subjective Well-Being Scales under the Presence of Acquiescent Response Style for the Racially and Ethnically Diverse Older Population in the United States. Surv Res Methods. 2020;14:417–37.

Leitgöb H, Seddig D, Asparouhov T, et al. Measurement invariance in the social sciences: Historical development, methodological challenges, state of the art, and future perspectives. Social Science Research. 2022:102805. doi:10.1016/j.ssresearch.2022.102805.

Li, Cheng-Hsien. 2016. “Confirmatory factor analysis with ordinal data: Comparing robust maximum likelihood and diagonally weighted least squares.” Behavior research methods 48(3):936–49. doi:10.3758/s13428-015-0619-7.

Lubke G, Muthén BO. Performance of Factor Mixture Models as a Function of Model Size, Covariate Effects, and Class-Specific Parameters. Structural Equation Modeling: A Multidisciplinary Journal. 2007;14(1):26–47. doi:10.1080/10705510709336735.

Marksteiner T, Kuger S, Klieme, E. The potential of anchoring vignettes to increase intercultural comparability of non-cognitive factors. Assess Educ Princ Policy Pract. 2019; doi:10.1080/0969594X.2018.1514367.

Meitinger, K. Necessary but Insufficient: Why Measurement Invariance Tests Need Online Probing as a Complementary Tool. Public Opin Q. 2017;81:447–72.

Meitinger K, Davidov E, Schmidt P, Braun M. Measurement Invariance: Testing for It and Explaining Why It is Absent. Surv Res Methods. 2020;14:345–49.

Menold N, Toepoel V. Do Different Devices Perform Equally Well with Different Numbers of Scale Points and Response Formats? A test of measurement invariance and reliability. Sociol Methods Res. 2022; doi:10.1177/00491241221077237.

Menold N, Tausch A. Measurement of Latent Variables with Different Rating Scales: Testing Reliability and Measurement Equivalence by Varying the Verbalization and Number of Categories. Sociol Methods Res. 2016;45:78–99.

Menold N, Kemper CJ. The impact of frequency rating scale formats on the measurement of latent variables in web surveys – An experimental investigation using a measure of affectivity as an example. Psihologija. 2015;48:4:431–49.

Meredith W. Measurement Invariance, Factor Analysis and Factorial Invariance. Psychometrika. 1993;58:525–43.

Meyer, B.D., Mok, W.K., Sullivan, J. X. Household Surveys in Crisis. Journal of Economic Perspectives 2015; 29 (4): 199–226

Millsap RE. Statistical approaches to measurement invariance. New York: NY; 2011.

Mirzoev T, Kane S. What is health systems responsiveness? Review of existing knowledge and proposed conceptual framework. BMJ Global Health. 2017;2.

Mõttus R, Allik J, Realo A, Pullmann H, Rossier J, Zecca G, Ah-Kion J, Amoussou-Yéye D, Bäckström M, Barkauskiene R, Barry O, Bhowon U, Björklund F, Bochaver A, Bochaver K, De Bruin GP, Cabrera HF, Chen SX, Church AT, Cissé DD, Dahourou D, Feng X, Guan Y, Hwang H-S, Idris F, Katigbak MS, Kuppens P, Kwiatkowska A, Laurinavicius A, Mastor KA, Matsumoto D, Riemann R, Schug J, Simpson B, Ng- Tseung C. Comparability of Self-Reported Conscientiousness Across 21 Countries. Europ J Pers. 2012;26:303–17.

Muthén BO. Beyond SEM: General Latent Variable Modeling. Behaviormetrika. 2002;29(1):81–117. doi:10.2333/bhmk.29.81.

Muthén BO, Asparouhov T. BSEM measurement invariance analysis. Mplus Web Notes, No. 17. 2013. http://www.statmodel.com/examples/webnotes/webnote17.pdf. Accessed 06. Apr 2022.

Muthén LK, Muthén BO. Mplus User’s Guide. Los Angeles, CA: Muthén & Muthén; 2014.

OECD. PISA 2012 Technical Report. OECD Publishing, Paris; 2014.

Paulhus DL. Measurement and control of response bias. In: Robinson JP, editor. Measures of social psychological attitudes: Vol. 1. Measures of personality and social psychological attitudes. San Diego: Academic Press; 1991. p. 17-59.

Rabe-Hesketh S, Skrondal A. Estimating CHOPIT models in GLLAMM: Political efficacy example from King, et al. (2002). 2002. http://www.gllamm.org/chopit.pdf. Accessed 06 Apr 2022.

Raftery AE. Bayesian Model Selection in Social Research. Sociol Method. 1995;25:111–63.

Raykov T, Marcoulides GA. A First Course in Structural Equation Modeling. Mahwah, NJ: Lawrence Erlbaum and Associates; 2016.

Raykov T, Marcoulides GA. Introduction to psychometric theory. New York: Taylor & Francis; 2011.

Rice N, Robone S, Smith PC. Vignettes and health systems responsiveness in cross-country comparative analyses. J R Stat Soc Ser A Stat Soc. 2012;175:337–69.

Roberts C, Sarrasin O, Ernst Stähli M. Investigating the Relative Impact of Different Sources of Measurement Non-Equivalence in Comparative Surveys. Surv Res Methods. 2020; doi:10.18148/srm/2020.v14i4.7416.

Salomon JA, Tandon A, Murray CJL. Comparability of self rated health: cross sectional multi-country survey using anchoring vignettes. BMJ. 2004;328(7434):258. doi:10.1136/bmj.37963.691632.44.

Stathopoulou T, Krajčeva E, Menold N, Dept S. Questionnaire design and translation for refugee populations: Lessons learnt from the REHEAL Study. Journal of Refugee Studies. 2019; doi:10.1093/jrs/fez045.

Tandon A, Murray CJL, Salomon JA, King G. Statistical models for enhancing cross-population comparability, In: Murray CJL, Evans DB, editors. Health Systems Performance Assessment: Debates, Methods and Empiricism. World Health Organization, Geneva. Taylor & Francis; 2003.

Tourangeau R, Rips LJ, Rasinski K. The Psychology of Survey Response. Cambridge: Cambridge University Press; 2000.

Valentine N, Prasad A, Rice N, Robone S, Chatterji S. Health systems responsiveness: a measure of the acceptability of health-care processes and systems from the user’s perspective. In: Smith PC, Mossialos E, Leatherman S, Papanicolas I, editors. Performance measurement for health system improvement. Cambridge: Cambridge University Press; 2009. p. 138–86.

van de Schoot R, Schmidt P, De Beuckelaer A, Lek K, Zondervan-Zwijnburg M Editorial: Measurement Invariance. Front Psychol.2015; doi:10.3389/fpsyg.2015.01064.

van Soest A, Delaney L, Harmon C, Kapteyn A, Smith JP. Validating the use of anchoring vignettes for the correction of response scale differences in subjective questions. J R Stat Soc Ser A Stat Soc. 2011; doi:10.1111/j.1467-985X.2011.00694.x.

van Vaerenbergh Y, Thomas TD. Response styles in survey research: A literature review of antecedents, consequences, and remedies. Int J Public Opin Res. 2013;25:195–217.

Wand, J., & King, G. (2007). Anchoring vignettes in R: A (different kind of) vignette. Retrieved from http://wand.stanford.edu/anchors/doc/anchors.pdf

Wolf EJ, Harrington KM, Clark SL. Sample Size Requirements for Structural Equation Models: An Evaluation of Power, Bias, and Solution Propriety. Educ Psychol Meas. 2013;73:913–34.

Wu AD, Li Z, Zumbo BD. Decoding the Meaning of Factorial Invariance and Updating the Practice of Multi-Group Confirmatory Factor Analysis: A Demonstration with TIMSS Data. Pract Assess Res Eval. 2007;12:1–26.

Yang Y, Harkness JA, Chin T-Y, Villar A. Response Styles and Culture. In: Harkness JA, Braun M, Edwards B, Johnson TP, Lyberg LE, Mohler PPh, Pennell B-E, Smith T-W, editors. Survey Methods in Multinational, Multiregional, and Multicultural Contexts. Hoboken, NJ: John Wiley & Sons; 2010. p. 203–223.

Zercher F, Schmidt P, Cieciuch J, Davidov E. The Comparability of the Universalism Value Over Time and Across Countries in the European Social Survey: Exact vs. Approximate Measurement Invariance. Front Psychol. 2015; doi:10.3389/fpsyg.2015.00733.

