## Supplemental Material for "Reducing Cross-Cultural Comparability Bias Obtained with Measurement Invariance Analysis by Means of Anchoring Vignettes in Heterogeneous Refugee Samples"

**Supplementary Material**

**Results of CHOPIT-Analysis**

While with the non-parametric approach we looked at the deviations in evaluations of vignettes, with the parametric approach we would like also to demonstrate how potential RC-DIF may impact evaluation of the variables under investigation. To demonstrate the potential effect of the RC-DIF on the results of the analyses, we compare the scores of cleanliness of the HSR as predicted by the regular ordered probit regression with the results of the CHOPIT analysis that used information on the potential RC-DIF form the vignettes. The results for two models are shown in Table 4. In the regular ordered probit model, the language indicator shows a significant and positive association with the rating of the quality of amenities of visited health care institutions, while the gender coefficient is not significant. The interpretation of these results would mean that Arabic-speaking refugees rate the cleanliness of facilities higher than English-speaking refugees, while there is no difference in the assessments of women and men. When controlling for RC-DIF with the help of the anchoring vignettes using CHOPIT analysis, these two associations are inverted. It is not the affiliation with language group, but with gender, that makes a difference in the assessment of cleanliness. Taking a closer look at the separate models that define the group specific thresholds by regressing them on the same covariates as the main model (bottom part of Table 4), we can see that the gender coefficient (.829) for the first vignette threshold – that is between responses “very bad” and “bad” – shows women more likely to choose the former category over the latter than men. Likewise, inspecting the results for the second threshold we find evidence for a divergence between refugees of different language groups (-1.765), which implies that speakers of the Arabic language refrain more often from using the “bad” instead of the “moderate” category. For the other thresholds, we can see the effects of age and health insurance card. These results again show the presence of RC-DIF, which also influences the effects of predictors on the dependent variable, as has been demonstrated in past research (e.g. Rice et al., 2012).

[insert Table 4 about here]

Table 4

Ordered Probit and CHOPIT Regression of Quality of Basic Amenities Ratings on Gender, Language, Age and Health Insurance Electronic Card

| | Ordered Probit | | CHOPIT | |
|---|---|---|---|---|
| | β | SE | β | SE |
| Female | 0.108 | (0.186) | 0.837* | (0.344) |
| Arabic speaking | 0.637*** | (0.191) | 0.493 | (0.277) |
| Age 25-29 | -0.239 | (0.208) | 0.0453 | (0.276) |
| Age 31-37 | -0.269 | (0.225) | 0.0258 | (0.420) |
| Age 38+ | -0.171 | (0.262) | 0.125 | (0.362) |
| Insurance Card | -0.0158 | (0.183) | 0.146 | (0.291) |
| Insurance Card - missing | -0.290 | (0.259) | -1.125* | (0.517) |
| Threshold 1 (very bad / bad) | | | | |
| Female | | | 0.829** | (0.284) |
| Arabic speaking | | | 0.224 | (0.201) |
| Age 25-29 | | | -0.234 | (0.208) |
| Age 31-37 | | | -0.392 | (0.308) |
| Age 38+ | | | -0.231 | (0.249) |
| Insurance Card | | | 0.513* | (0.253) |
| Insurance Card - missing | | | -0.712 | (0.411) |
| Constant | -1.698*** | (0.224) | -1.670*** | (0.273) |
| Threshold 2 (bad / moderate) | | | | |
| Female | | | -0.137 | (0.425) |
| Arabic speaking | | | -1.765* | (0.724) |
| Age 25-29 | | | 0.839** | (0.323) |
| Age 31-37 | | | 0.677 | (0.477) |
| Age 38+ | | | -0.219 | (0.822) |

| | | | | |
|---|---|---|---|---|
| Insurance Card | | | -0.270 | (0.328) |
| Insurance Card - missing | | | -0.378 | (0.616) |
| Constant | -1.294*** | (0.203) | -0.816** | (0.315) |
| Threshold 3 (moderate / good) | | | | |
| Female | | | 0.0298 | (0.421) |
| Arabic speaking | | | 0.557 | (0.351) |
| Age 25-29 | | | -0.162 | (0.384) |
| Age 31-37 | | | 0.267 | (0.539) |
| Age 38+ | | | 0.209 | (0.459) |
| Insurance Card | | | -0.493 | (0.407) |
| Insurance Card - missing | | | 0.442 | (0.462) |
| Constant | -0.923*** | (0.193) | -1.122*** | (0.316) |
| Threshold 4 (good / very good) | | | | |
| Female | | | -0.0912 | (0.238) |
| Arabic speaking | | | -0.214 | (0.209) |
| Age 25-29 | | | 0.477* | (0.238) |
| Age 31-37 | | | 0.681* | (0.272) |
| Age 38+ | | | 0.749** | (0.263) |
| Insurance Card | | | -0.145 | (0.210) |
| Insurance Card - missing | | | -0.239 | (0.342) |
| Constant | 0.0354 | (0.186) | -0.310 | (0.209) |
